# Understanding the key determinants of an HPV therapeutic vaccine: a modeling analysis

**DOI:** 10.1101/2023.12.04.23299403

**Authors:** Jamie A. Cohen, Robyn M. Stuart, Serin Lee, Daniel J. Klein, Cliff C. Kerr, Darcy W. Rao, Haina Shin, Sharon L. Achilles

**Affiliations:** Institute for Disease Modeling, Global Health Division, Bill & Melinda Gates Foundation, Seattle, Washington, USA; Data & Insights, Gender Equality Division, Bill & Melinda Gates Foundation, Seattle, Washington, USA; Department of Industrial & Systems Engineering, University of Washington, Seattle, Washington, USA; Reproductive Health Technologies, Global Health Division, Bill & Melinda Gates Foundation, Seattle, Washington, USA

## Abstract

Despite incredibly effective tools to prevent HPV infection and treat precancerous lesions, the scale-up of existing interventions in most low and middle-income countries has been slow, leaving a residual burden of invasive cervical cancer that will persist for decades. An HPV therapeutic vaccine may overcome some of the scalability and infrastructure challenges of traditional screening and treatment programs, though its potential public health value depends upon its characteristics, delivery strategy, and the underlying immunity of the population on which it would act. This analysis uses HPVsim, an open-access agent-based simulation framework, to evaluate the impact of a range of potential HPV therapeutic vaccines with varying scale-up of existing preventive interventions in nine high-burden low- and middle-income countries (LMICs). For each setting, the model is populated with context-specific demographic and behavioral data, and calibrated to fit estimates of HPV and cervical disease by age. We find that an HPV therapeutic vaccine that clears 90% of virus and regresses 50% of high-grade lesions, reaching 70 percent of 35-45 year old women starting in 2030, could avert 1.2-2.2 million incident cases of cervical cancer, 500,000-1.2 million cervical cancer deaths and 20-40 million disability adjusted life years (DALYs) in the modeled high-burden LMICs over 30 years. The size of the impact is sensitive to rates of background intervention scale-up and the characteristics of the vaccine, including ability to establish long-lasting immune memory.

## 1. Introduction

Cervical cancer elimination, defined as reaching and maintaining an incidence rate of below 4 per 100,000 women, is an important global public health goal, with the potential to avert over 300,000 deaths globally per year. Modeling has estimated that cervical cancer elimination can be achieved through rapid adoption and scale-up of prophylactic vaccination to 90% of young girls, screening 70% of women between ages 30 and 50 years for pre-cancer, and treatment of 90% of detected pre-cancers and cancers. However, most low and lower-middle income countries (LMICs) remain far from reaching these 90-70-90 WHO elimination targets (*1–3*). As of 2019, only 40% of LMICs had introduced HPV vaccination into their national programs and only 15% of girls in the target age for HPV vaccination had received two doses (*4*). Only 10% of eligible women have ever received screening in LMICs, compared to 84% of women in high-income countries (*5*).

HPV prophylactic vaccines have the potential to significantly reduce the burden of cervical cancer by preventing the acquisition of high-risk HPV. The main targeted population for HPV prophylactic vaccines is girls aged 9-14 years. Despite the significant promise of this intervention, scale-up has been challenged by vaccine hesitancy, insufficient supply, and a scarcity of delivery platforms for this specific age group, which is not otherwise traditionally reached by vaccination programs. Additionally, even with optimistic scale-up of prophylactic vaccination, reductions in cervical cancer burden will not accrue until several decades into the future, and will not be realized by women who missed prophylactic vaccination prior to HPV infection and remain at risk of persistent oncogenic infection that can progress to cervical cancer.

For unvaccinated women, screening is the most effective tool for early identification of pre-cancerous lesions with subsequent treatment that prevents progression (*6, 7*). HPV DNA testing has transformed the ability to detect high-risk infection with a high degree of sensitivity, and thermal ablation has enabled low-cost treatment of precancerous lesions (*8*– *12*). Screening and treatment programs remain expensive and challenging to scale-up in many LMIC geographies (*13–15*). The high cost of HPV DNA tests often leads to reliance on visual inspection with acetic acid (VIA), which is known to have both low sensitivity and variable performance, resulting in imprecise treatment decisions. Further, since screening and treatment are not typically done in a single visit, there is significant loss to follow-up (*16–18*).

Several therapeutic vaccines have been tested, with varying success. Vaccines targeting early stage invasive cervical cancer and high-grade lesions have been tested in clinical trials, several products that work to clear infection have reached phase 1/2a trials, and one candidate targeting productive infection is in early stages of discovery (*19–21*). These HPV therapeutic vaccines have the potential to meet the needs of women who have been missed by prophylactic vaccination and may circumvent some of the scalability and infrastructure challenges of traditional screening and treatment programs. However, the impact of a therapeutic vaccine depends upon how quickly it can be developed and delivered, the characteristics of the product and its mechanism of action, and the degree and timing of scale-up of prophylactic vaccination and screening and treatment.

Resources devoted to discovery and development of such a product have a high opportunity cost, as they could alternatively support the scale-up of existing interventions. Therefore, understanding the full public health value of an HPV therapeutic vaccine under a range of plausible future scenarios and against scientific uncertainty is critical. To date, there has been one published study (*22*) that uses dynamic modeling to evaluate the potential impact of an HPV therapeutic vaccine in Uganda. The present study builds from that analysis by exploring the impact of potential HPV therapeutic vaccine candidates with varying efficacy for virologic clearance and regression of high-grade pre-cancer, as well as its ability to induce long-term memory. We quantify the incremental impact of potential HPV TxV candidates in nine high-burden low and middle-income countries with a large burden of disease and low coverage of screening and treatment.

## 2. Methods

### 2.1 Methods overview

Using HPVsim, an open-access agent-based simulation framework for estimating the burden of cervical cancer, we calibrated nine models to epidemiologic data in India, Indonesia, Bangladesh, Myanmar, Ethiopia, Nigeria, Democratic Republic of Congo, Uganda, and Tanzania. These countries were chosen based upon their cumulative disease burden and current membership in GAVI. The HPVsim platform is described in Stuart et al. (*23*) and in the supplemental appendix. Briefly, HPVsim simulates individual agents who age over time, participate in structured sexual networks, and can acquire genotype-specific HPV infection. HPV can clear and lead to seroconversion and associated B- and T-cell immunity which protects against subsequent type-specific HPV acquisition and progression, or persist. Persistent high-risk infections have an accumulating risk of progression to invasive cervical cancer over time in the absence of an intervention.

In the remainder of this section, we provide details on how the models were calibrated (Section 2.1) and the scenarios and sensitivity analyses that we ran (Section 2.2). All analysis code is publicly available online, as is the [HPVsim](http://hpvsim.org/) model itself.

### 2.1. Model calibration and validation

We calibrated nine models by adjusting a subset of the model’s input parameters in order to achieve a good match to estimates on cervical cancer cases by age and, where data was available, the distribution of HPV types among women with precancerous lesions and invasive cervical cancer. In all countries, we prophylactic vaccination and screening and treatment are low or absent; we make a simplifying assumption of no prophylactic vaccination or screening and treatment programs prior to 2020. Details and results of the calibration procedure can be found in section 1.3 of the supplemental appendix.

### 2.2. Scenario analyses

The impact of a therapeutic vaccine is highly dependent on the relative scaleup of screening for pre-cancer and treatment of pre-cancer and invasive cervical cancer. We assume that scale-up of TxV is unlikely in the absence of PxV, such that all modeled scenarios include 90% coverage of PxV. Scale-up of routine screening and treatment (S&T), however, has been more challenging and may continue to be delayed or absent, and we evaluate a range of scenarios of S&T scale-up (Table 1). For each scenario, we first estimate residual burden until 2060 in the absence of therapeutic vaccination to formulate a baseline counterfactual against which to evaluate a therapeutic vaccine. We then run the scenarios with TxV and compute the number of cervical cancer cases, deaths, and DALYs averted from 2030 to 2060.

**Table 1.** Prophylactic vaccine and S&T scenarios considered. Lifetime coverage of S&T has been estimated at less than 5% in all settings, though there might be significant regional variation.

|  | Prophylactic vaccination | Screening & treatment (S&T) |
| --- | --- | --- |
| **Scenario 1** | 90% coverage | 0% screen and treatment coverage |
| **Scenario 2** | 90% coverage | 35% screen coverage, 70% treatment coverage |
| **Scenario 3** | 90% coverage | 70% coverage, 90% treatment coverage |

We assume the nonavalent HPV prophylactic vaccination is introduced in each country in 2025 and administered to 9-14 year old girls, reaching 90 percent coverage level immediately. Vaccination is assumed to generate high and long-lasting type-specific immunity against infection. The level of protection is drawn per person at the time of vaccination and applied per sex act, meaning the vaccine is modeled as a “leaky vaccine” (*24*). Screening is similarly introduced in 2025 and consists of an HPV DNA test for women twice in their lifetime between ages 30 and 50, reaching the target coverage level immediately. Women who receive a positive result are eligible for ablative or excisional treatment, with 81 and 93 percent efficacy in regressing pre-cancerous lesions respectively.

On top of these alternative prophylactic vaccination and S&T scenario, we evaluate a range of scenarios varying the characteristics and delivery of an HPV therapeutic vaccine (see Table 2). We consider targeting 30, 35, or 40-year-old women for routine delivery starting in 2030, and conduct a one-time campaign in 2030 reaching women between the target age and the next 10 birth cohorts. For all scenarios, we assume the product would require two doses administered 3 months apart, and that 70% of eligible women are reached with the first dose and that 20% of women who receive their first dose will not return for their second dose. We assume the first dose will provide half the efficacy of the completed regimen.

**Table 2.** Therapeutic vaccine assumptions in base case and values considered in sensitivity analyses. Effectiveness values (e.g.., 90/0) represent full effectiveness of a completed 2-dose regimen in clearing HPV infection as the first value (i.e., 90%) and regressing pre-cancer as the second (i.e., 0%). TxV: HPV therapeutic vaccine.

|  | Base case | Sensitivity |
| --- | --- | --- |
| **TxV effectiveness** | 90/0, 70/30, 50/50, 90/50 | N/A |
| **Target age of routine administration** | 30, 35, 40 | N/A |
| **Age of catch-up campaign (in 1<sup>st</sup> year of introduction)** | Target age – target age + 10 | N/A |
| **TxV immune memory** | Maximum of efficacy against virus and lesion<br>Waning: lifelong | No durable memory |
| **Coverage** | 70% first-dose coverage, 20% LTFU between doses | N/A |
| **Doses** | 2-doses, 3 months apart | N/A |
| **Introduction year** | 2030 | 2035 |
| **Cross-protection** | None | 50% cross protection against all other high-risk genotypes |

In the absence of trial data on effectiveness of a therapeutic vaccine product, we conducted robust exploration around therapeutic vaccine properties, varying effectiveness against productive infections from 50 to 100% and against high-grade lesions from 0 to 50%. An upper-bound of 50% effectiveness at regressing high-grade lesions was imposed based upon modest results of previous therapeutic vaccines that have targeted high-grade lesions and cervical cancer and the expectation that the microenvironment would be less conducive for stimulating a response that would effectively regress high-grade lesions (*25*).

The relative impact of the therapeutic vaccine depends on how effective it is at both virological clearance and lesion regression. It is expected that this vaccine will work by inducing a CD8 T-cell response that will clear active infection and/or lesion (*26*). The magnitude, breadth, and duration of that immune response will depend upon the immunogen design of the vaccine, existing HPV-specific T-cell repertoire, and mode of delivery.

In the base case, we assume the therapeutic vaccine will provide no cross protection against other high risk HPV types. We assume that the therapeutic vaccine acts on prevalent infection and/or lesion, based upon its attributes, and induces the same level of long-lasting immunity to reduce the severity of future infections and reactivations as the effectiveness generated by each dose of the therapeutic vaccine.

The relative impact of the therapeutic vaccine depends on how effective it is at both virological clearance and lesion regression. It is expected that this vaccine will work by inducing a CD8 T-cell response that will clear active infection and/or lesion (*26*). The magnitude, breadth, and duration of that immune response will depend upon the immunogen design of the vaccine, existing HPV-specific T-cell repertoire, and mode of delivery. Given this uncertainty, we consider the impact of immune memory and effectiveness of such a product on its potential public health impact, co-varying the level of immunity against disease from a future infection and the vaccine’s effectiveness against prevalent virus and lesion.

We run each scenario multiple times using the best fitting parameter set and 5 different random seeds and present the mean and ± two times standard deviation of the results.

## 3. Results

### 3.1 Residual burden

We find a large residual burden of cervical cancer over the next 35 years for all scale-up scenarios without TxV for all countries modeled, with the majority of burden driven by India. Even in the context of optimistic prophylactic vaccination and S&T scale-up (achieving the 90-70-90 WHO elimination targets) in all 9 countries, the model estimates over 200,000 new cases of cervical cancer each year (see Figure 1).

**Figure 1.**
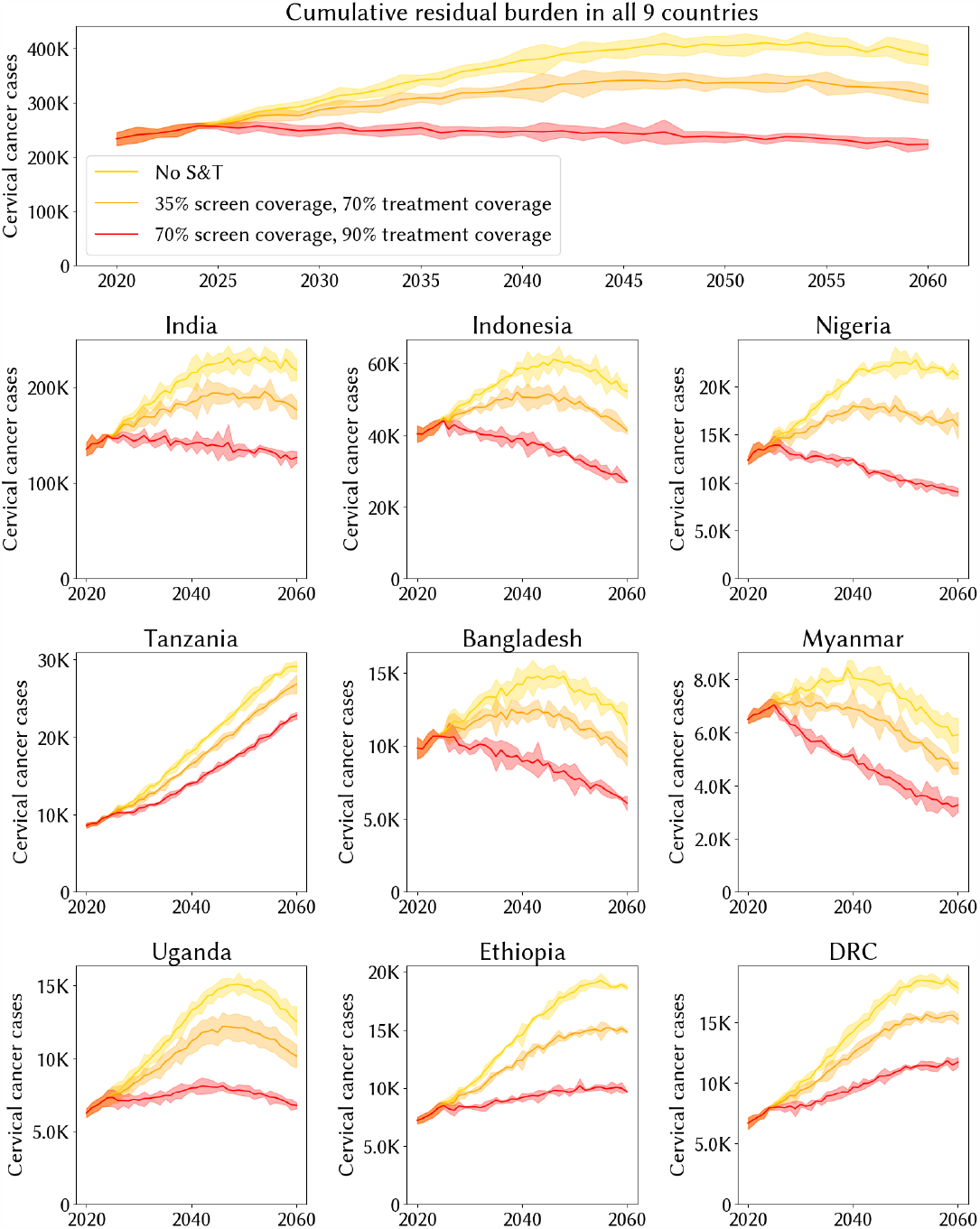
Residual annual burden of incident cervical cancers in nine high-burden LMICs from 2020 until 2060, assuming different rates of S&T coverage scale-up. S&T: screening and treatment. Top panel plots the cumulative residual burden across all nine countries.

### 3.2 Therapeutic vaccine impact

We first consider the relationship between age of targeted routine TxV delivery, TxV characteristics, and background scale-up of PxV and S&T. The impact of TxV on cervical cancer cases, deaths, and DALYs is greatest for a product that clears virus with 90% effectiveness and regresses CIN2+ with 50% effectiveness (Figure 2). With background screening and treatment coverage and ages held fixed, TxV effectiveness against pre-cancerous lesion yields the biggest impact over the short term, while TxV with effectiveness in clearing virus has delayed impact.

**Figure 2.**
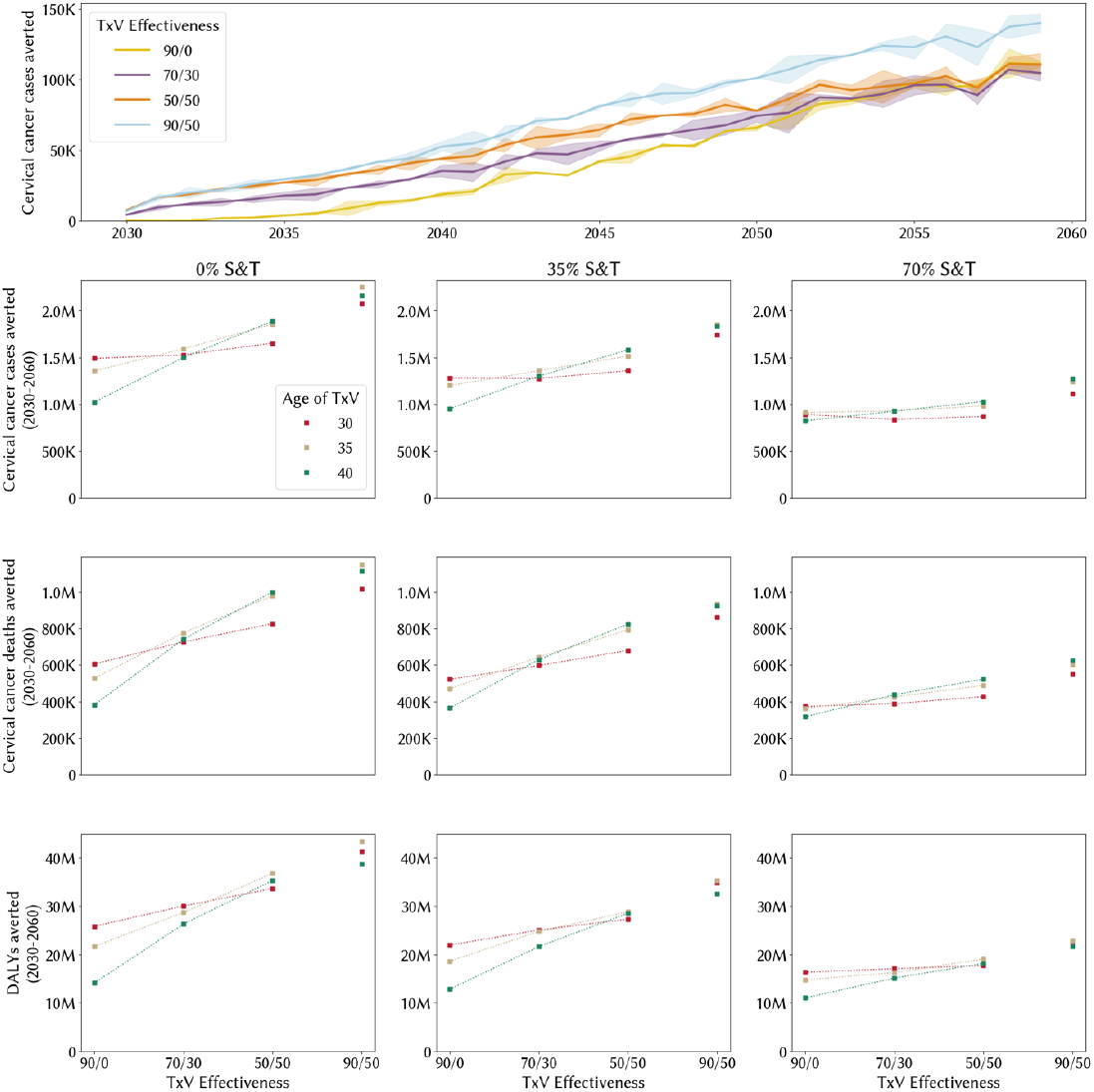
Health impact of an HPV therapeutic vaccine summed across nine high-burden LMICs based upon effectiveness, background intervention scale-up, and age of administration. Effectiveness values (i.e. 90/0) represent full effectiveness of a completed 2-dose HPV therapeutic vaccine regimen in clearing HPV infection first (i.e. 90%) and regressing high grade pre-cancer second (i.e. 0%). Time series plotted for the background intervention scenario of 90% PxV coverage and no S&T scale-up only and for a target TxV age of 35.

**Figure 3.**
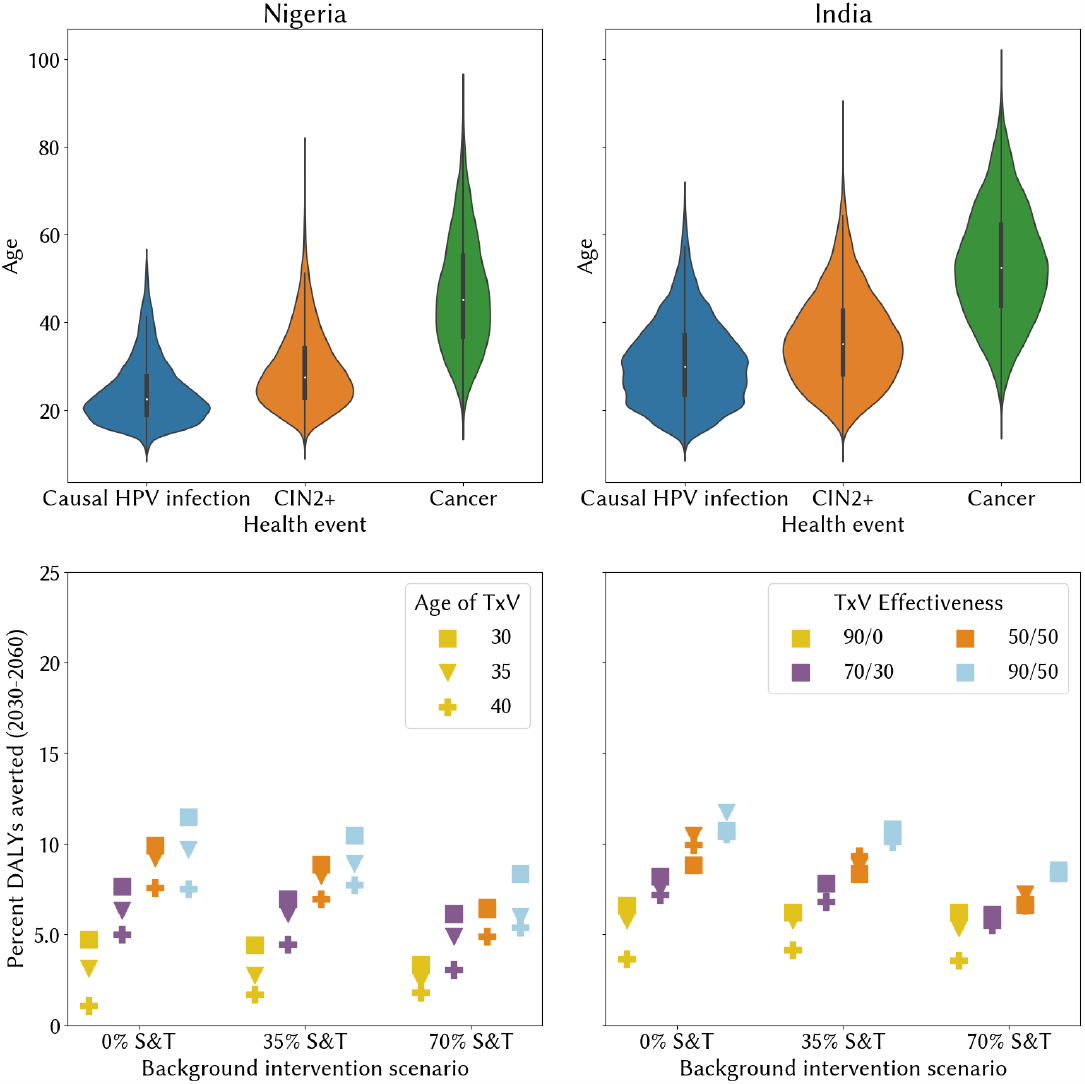
First row shows the distribution of age of causal HPV infection, CIN2+, and cancer in Nigeria and India. The second row shows DALYs averted due to administration of an HPV therapeutic vaccine in Nigeria and India based upon effectiveness, background intervention scale-up, and age of administration.

For all three metrics, the health impact is greatest for a TxV candidate that clears virus with 90% effectiveness and regresses pre-cancerous lesion with 50% effectiveness and in the absence of screening and treatment scale-up. The time series plot in Figure 2 shows the timescales of impact holding fixed the level of background screening and treatment coverage and age of administration, whereby effectiveness against pre-cancerous lesion yields the biggest impact over the short term, while effectiveness against virus has a delayed impact. TxV candidates with 50% effectiveness against precancerous lesions have the highest early impact, but at 7 years post-introduction the impact of the product that also has 50% effectiveness for viral clearance (orange line in Figure 2) begins to level off compared to the candidate with 90% effectiveness for viral clearance (blue line). The impact of a candidate with 90% viral clearance and no impact on precancerous lesions (yellow line) converges with candidates with moderate impact on precancerous lesions and lower viral clearance (orange and purple lines) approximately 25 years post-introduction.

When we consider the trade-off between effectiveness against virus and high-grade pre-cancer alongside age of administration, we find the age target that maximizes cancer cases and deaths averted switches from 30 to 35 to 40 as lesion regression effectiveness grows from 0 to 30 to 50%. Irrespective of screening and treatment scale-up, there is more value to regressing high grade lesions for older women than younger women. This can be visualized by the slope of the green lines in Figure 2, representing administration to women age 40, always being steeper than the tan (age 35) and red (age 30) lines. The slope of all three lines flattens as screening and treatment coverage increases from 0 to 35 to 70%, indicating that there is a smaller difference in age of administration for different candidates.

When using DALYs as a metric of health impact, targeting women age 30 remains the most effective until the effectiveness against high-grade lesions reaches 50%. For all other TxV candidates and levels of screening and treatment scale-up, targeting age 35 maximizes DALYs averted.

We then explored the extent to which these findings are consistent across settings, hypothesizing that the biggest differences would be seen in settings with different epidemiologic age profiles. To do this, we compared the health impact of TxV in Nigeria, with a median age of cancer of 45, to India, with a median age of cancer of 55, representative of both extreme ends of the spectrum within our nine modeled countries.

The age that maximizes DALYs averted is equivalent or younger in Nigeria than in India for every candidate profile and background level of screening and treatment coverage. In Nigeria, it is always optimal to target 30 year-old women for routine administration of TxV, whereas the optimal age in India depends upon the TxV effectiveness against virus and lesion as well as background level of screening and treatment coverage. In both settings, the overall health impact increases monotonically with more effectiveness against pre-cancerous lesion for each age target.

To understand the additional important levers that might influence the value of a TxV, we considered the inclusion of 50% cross-protection against other high-risk genotypes, a 5-year delay in the introduction of such a TxV candidate, and the absence of durable immune protection. We considered these three sensitivity analyses for a TxV candidate with the most optimistic effectiveness profile, with delivery targeting 30, 35, or 40 year old women. Figure 4 shows the associated impact on DALYs averted compared to baseline for each sensitivity analyses, both cumulatively and individually per country.

**Figure 4.**
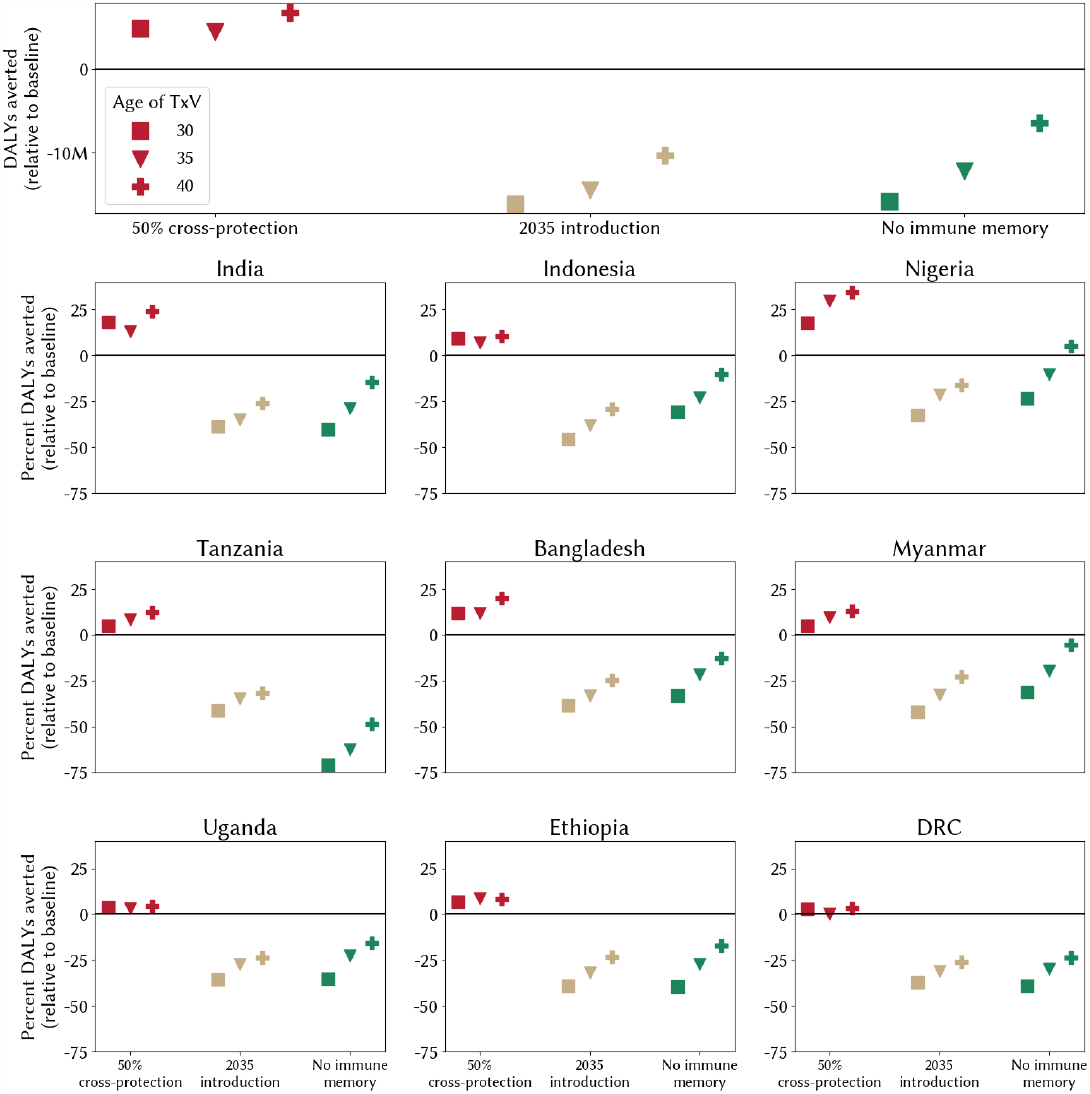
Sensitivity analysis of cross-protection for non-16/18 high-risk genotypes, delaying TxV introduction by 5 years, and absence of durable immune memory. All findings presented assume no screening and treatment.

Adding in 50% cross-protection would avert an additional 5-7 million DALYs over 30 years compared to the same TxV candidate with no cross protection, with significant variation across countries. Cross protection would increase DALYs averted by up to 28% in Nigeria, where nearly 40 percent of cervical cancers are attributable to non-16/18 HPV types, whereas it would have a negligible impact on DALYs averted in DRC, where less than 5% of cervical cancers are attributable to non-16/18 HPV types. In contrast, delaying TxV introduction by 5 years would result in over 10 million fewer DALYs averted, with significantly less variation by country.

Most notable is the loss in health impact associated with a TxV candidate without durable immune memory, with both tremendous variation by age of delivery and by country. For all countries, there is a larger loss in DALYs averted when TxV is administered to 30 year-olds than 35 or 40 year-olds, reflecting the fact that women at younger ages might not yet have acquired the infection/lesion that would progress to invasive cervical cancer in the absence of treatment. More striking is the difference across countries, with the lack of durable immune memory having the smallest impact on DALYs averted in Nigeria (0-25% fewer than with immune memory) and the largest impact in Tanzania (50-75% fewer). These differences reflect associated differences in the age distribution of infection and cancer, with Nigeria having a nearly 15-year younger age of causal HPV infection than Tanzania.

## 4. Discussion

Using a robust dynamic simulation model fit to 9 high-burden LMICs, we find that HPV TxV can have a significant impact on burden of disease, and that this impact depends strongly on scale-up of screening and treatment and TxV candidate characteristics. Importantly, we find that a TxV that has some effectiveness at regression of high-grade lesions accelerates and increases burden reduction relative to one that only clears productive infection. This mirrors the delayed impact of prophylactic vaccines due to the age of administration and mechanism of impact and reveals the cost of a therapeutic vaccine without significant action against pre-cancerous lesions – it leaves avertable DALYs on the table. Putting these results in context, we find that, even with the WHO 90-70-90 targets for PxV and S&T and in a short-term time horizon evaluation, an HPV therapeutic vaccine could avert over 20 million DALYs in nine high-burden LMICs over 30 years. Given that product development will take place in parallel with ongoing efforts to scale up existing cervical cancer programming, this analysis reveals that a therapeutic vaccine would likely have tremendous benefit regardless of the level of progress made with these parallel efforts.

This study finds that an HPV therapeutic vaccine might avert 1-2 million incident cases of cervical cancer, 400,000 - 1.2 million deaths from cervical cancer, and 20-40 million DALYs in nine high-burden LMICs over 30 years, depending upon the TxV candidate characteristics and level of intervention scale-up. The findings in this analysis are aligned with the results of the Spencer et al. analysis (*22*), which found 139,000 fewer incident cervical cancers associated with an HPV therapeutic vaccine delivered to women in Uganda (compared to approximately 120,000 incident cases averted in Uganda in the current analysis, see Figure A16 for details).

In addition to uncertainty in the scale-up of existing interventions and characteristics of a potential TxV candidate, there also exists real uncertainty and variation in many of the mechanisms and processes being modeled, including in behavior and natural history. This variation might reveal contexts in which an HPV therapeutic vaccine would have more impact relative to other contexts or might target different age groups to maximize value, due to factors such as different patterns of sexual behavior. Our analysis attempted to draw out some of this variation by comparing across nine high-burden LMICs with significant differences in epidemiologic profiles. Future work in this area should consider development of sub-national models to capture real variation in epidemiology seen across and within settings (*27*) and should be applied to other settings with varied epidemiologic and demographic patterns to test generalizability.

## Data Availability

All data produced are available online at https://github.com/amath-idm/hpvsim_txvx_analyses

## Acknowledgments

We would like to thank Celina Schocken, Holger Kanzler, Ru Cheng, Sami Gottlieb, and Holly Prudden for their editorial and scientific review of this work.

