## Supplemental appendix for "Understanding the key determinants of an HPV therapeutic vaccine: a modeling analysis"

### 1 Model Overview

We created a model of HPV transmission and progression in 9 separate high-burden, low- and middle-income countries using the HPVsim open-source modeling framework (*1*). Elsewhere, we describe how the HPVsim software can capture demographics and simulate HPV transmission over realistic sexual networks (*2*), and in Section 1.1 we provide more details on how this was done to represent the 9 countries modeled in this analysis. Section 1.2 describes the natural history model that was used for this analysis. Section 1.3 describes the model calibration process and results.

#### 1.1 Adaptation of HPVsim

This analysis modeled the countries of India, Indonesia, Bangladesh, Myanmar, Ethiopia, Nigeria, Democratic Republic of Congo, Uganda, and Tanzania from 1950 until 2050 and relied upon HPVsim’s automated demographic features to initialize and update the population with births, deaths of causes other than cervical cancer, and migration. The model’s demographic components rely on population estimates, birth rates by time, and death rates by time, age, and sex from the UN’s World Population Projections (*24*). To capture HPV transmission, we include four genotypes: 16, 18, a pooled combination of all non-16/18 high-risk types included in the nonavalent prophylactic vaccine (31, 33, 45, 52, and 58), and a pooled combination of all other high-risk types (including types 35, 56, and 59).

The model uses a female-driven pair formation algorithm, where women are eligible for a new sexual partnership if they have reached or exceeded their assigned age of sexual debut and are under-partnered (defined as fewer sexual partners than are desired). A suitable male partner is selected for all eligible women based upon age-mixing matrices. We model three types of sexual relationships – marital, casual, and one-off – defined by their duration, condom usage, propensity for concurrency, and age mixing. These sexual network parameters do not change temporally in the model. We conducted a pre-calibration step of modifying the participation rate by age and sex to fit observed sexual behavior data, though are limited by information on rates of marriage and concurrency.

We adapted HPVsim’s built-in sexual network algorithm to roughly fit demographic health surveys data on median age of coitarche (first sexual experience), probability of marriage by age, and percent of women reporting having had sex with at least one non-marital partner. In the absence of robust and population-representative sexual behavior data, we calibrated the probability of men and women participating in extra-marital relationships and the number of concurrent casual partnerships for men and women.

#### 1.2 Natural history

HPVsim contains an embedded model of the natural history of HPV and cervical disease, which has been adapted to model cervical cancer epidemiology in 9 high-burden LMICs. Briefly, HPV infections are assigned a genotype-specific duration which can be moderated by factors such as immunity and age. This duration is then used to determine the likelihood of progression to pre-cancerous lesions. Among women whose infection progresses to pre-cancer, a duration of pre-cancer is sampled and used to determine the likelihood of progression to invasive cervical cancer. We held fixed the default duration of HPV infection prior to progression or clearance and the probability of progression to pre-cancer, both of which were parameterized based upon data from 7 years of follow up of the control arm of the Costa Rica Vaccine Trial (*3*); and the level of neutralizing immunity generated after seroconversion, informed by serologic assays of patients in the Guanacaste Natural History Study (*4*).

We then calibrated key components of the HPV natural history to fit cervical cancer cases by age and HPV genotype distribution in pre-cancer and invasive cervical cancer in India. Specifically, we calibrated the genotype-specific duration of pre-cancerous lesions prior to either invasive cervical cancer or regression; and parameters of a function mapping duration of pre-cancerous lesion to probability of developing invasive cervical cancer.

#### 1.3 Calibration

##### 1.3.1 Calibration methods

All calibration targets were derived from Globocan 2020 estimates and HPV Information Centre compiled meta-analyses (*5*). A description and search range for each of the 26 calibrated parameters is shown in Table A1. We use Optuna (*6*), a Bayesian hyperparameter optimization algorithm, to search across these parameter ranges to find solutions that fit the data well. We quantify goodness-of-fit as the sum of normalized absolute differences (SNAD) between the model’s outputs and the data, scored as Equation 1, and we aim to find parameter combinations that minimize the SNAD (Equation 2).

${GOF}_{i}=\sum_{j}^{n_{i}} model_{i,j}-data_{i,j}$	(1)

$SNAD = \sum_{i}^{L} \frac{GOF_{i}}{\max_{j \in\left( 1, \ldots, n_{i} \right)} \left( data_{i,j} \right)\cdot n_{i}}$	(2)

for $i$ in L calibration targets, where each target is composed of a series of $n_{i}$ data points across age groups or genotypes. We ran 8,000 trials per country and present the top 50 best-fitting parameter sets for each country.

| **Parameter name** | **Description** | **Lower, upper** |
| --- | --- | --- |
| beta | Per-act probability of HPV transmission | 0.02, 0.5 |
| age risk | Factor by which severity of infection increases at older ages  Age at which severity of infection increases | 1, 4  30, 50 |
| own_imm_hr | Degree of protection against subsequent infection of the same group for ohr and hi5 genotype groups | 0.25, 1 |
| dur_cin | Duration of pre-cancerous lesion prior to regression or progression (lognormal distribution) | par1=3,12 par2=10, 25 |
| cancer_fn | ‘aggressiveness’ parameter describing how quickly dysplasia progresses to a potentially oncogenic state | 15, 30 |
| f_cross_layer  m_cross_layer | Probability of men and women to participate in extra-marital partnerships | F: 0.05, 0.5  M: 0.1, 0.7 |
| f_partners  m_partners | Number of concurrent casual partners (poisson distribution) | F: 0.1, 0.6  M: 0.1, 0.6 |

**Table A1.** Parameters varied in calibration, including a description and the lower and upper bounds that were searched in the sampler.

##### 1.3.2 Calibration results

We were able to achieve a good fit to all target data for all 9 countries, capturing both the age trend of cervical cancer cases as well as HPV type distribution in pre-cancer and invasive cervical cancer (see Figure A1 for fit to cancers by age for all 9 countries and Figure A2-A10 for the individual fit to target data per country). Figure A11 and A12 show the implied natural history of the best-fitting parameter sets of this calibration used for the analysis.

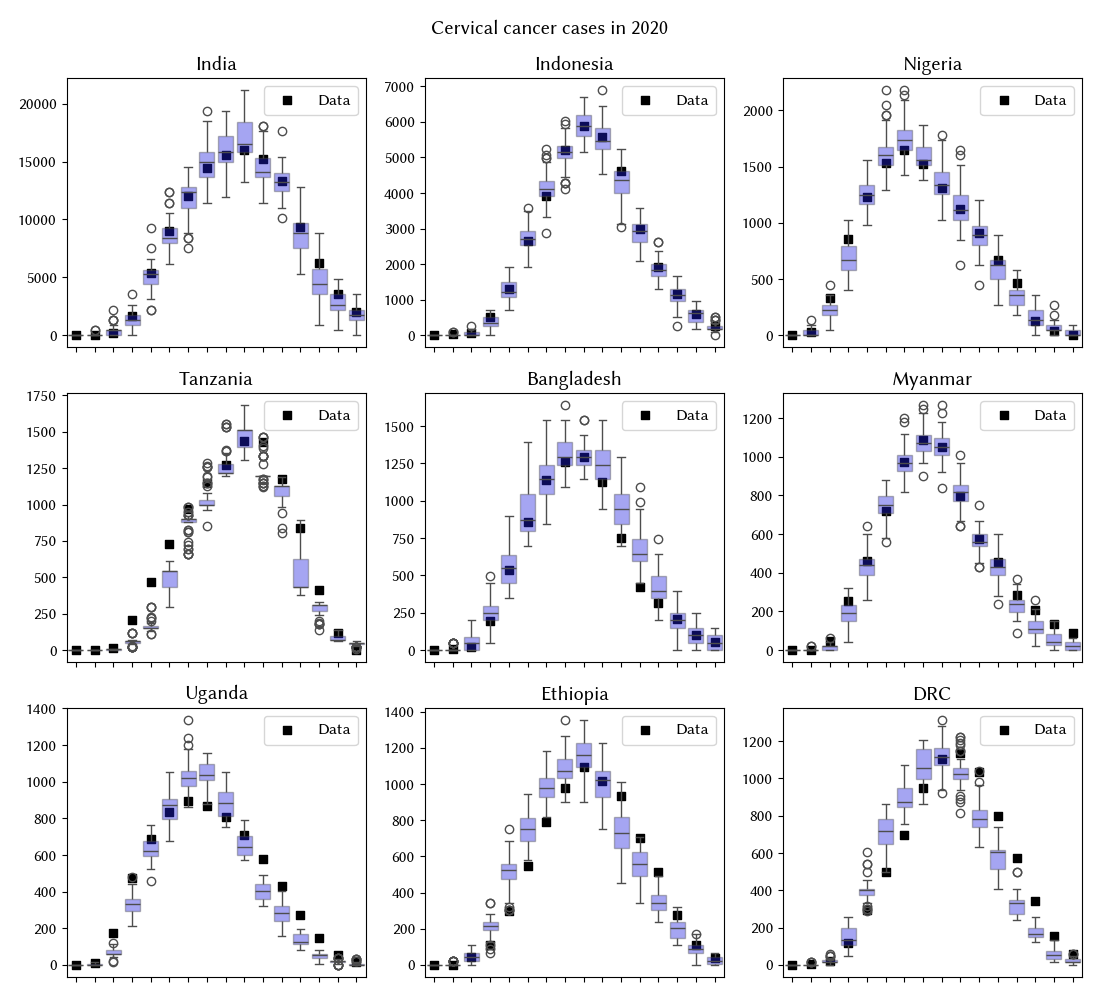

**Figure A1.** Calibration results for cancers by age across the 9 countries. Model output for the top 100 parameter sets is plotted alongside corresponding target data.

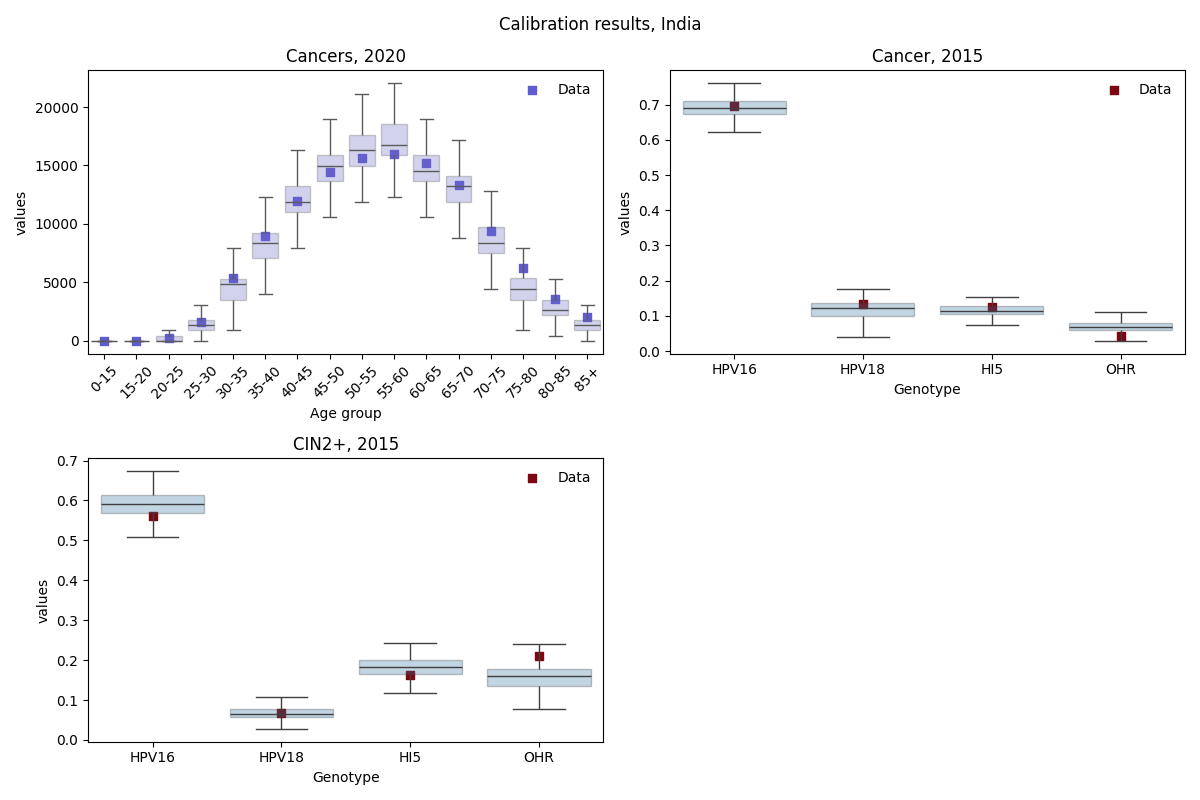

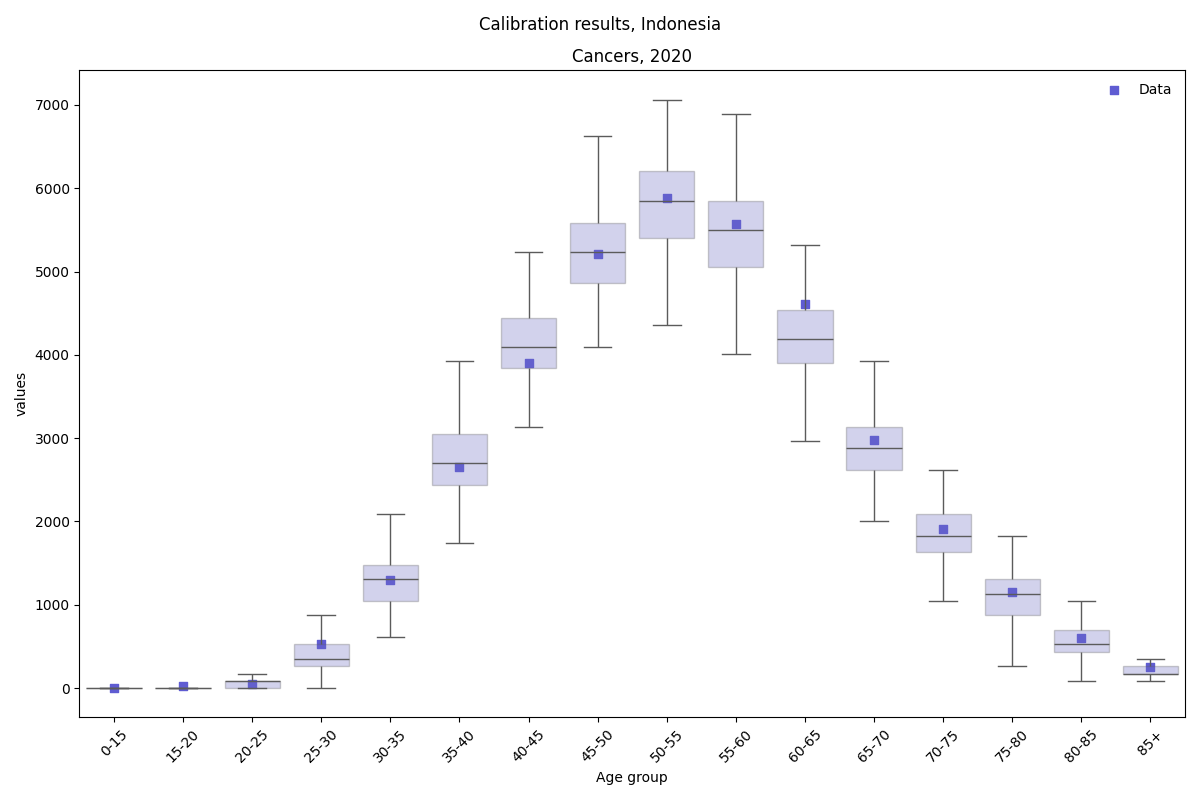

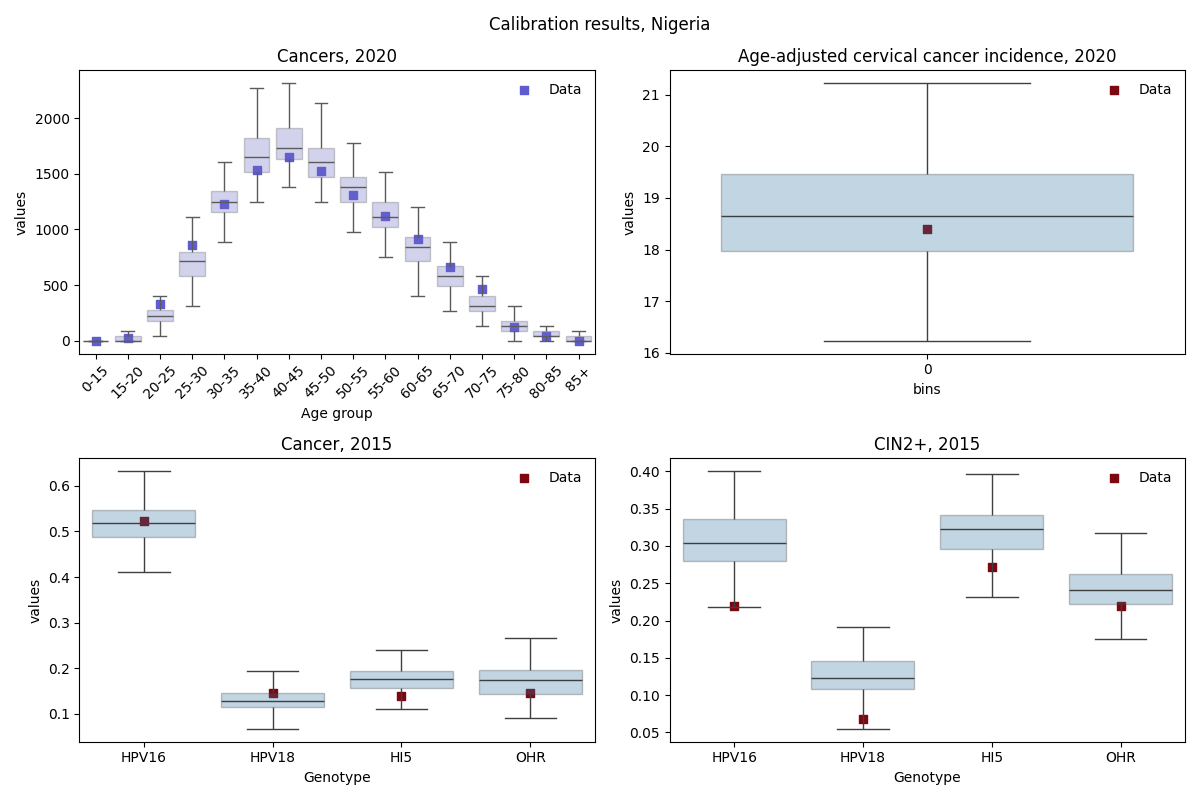

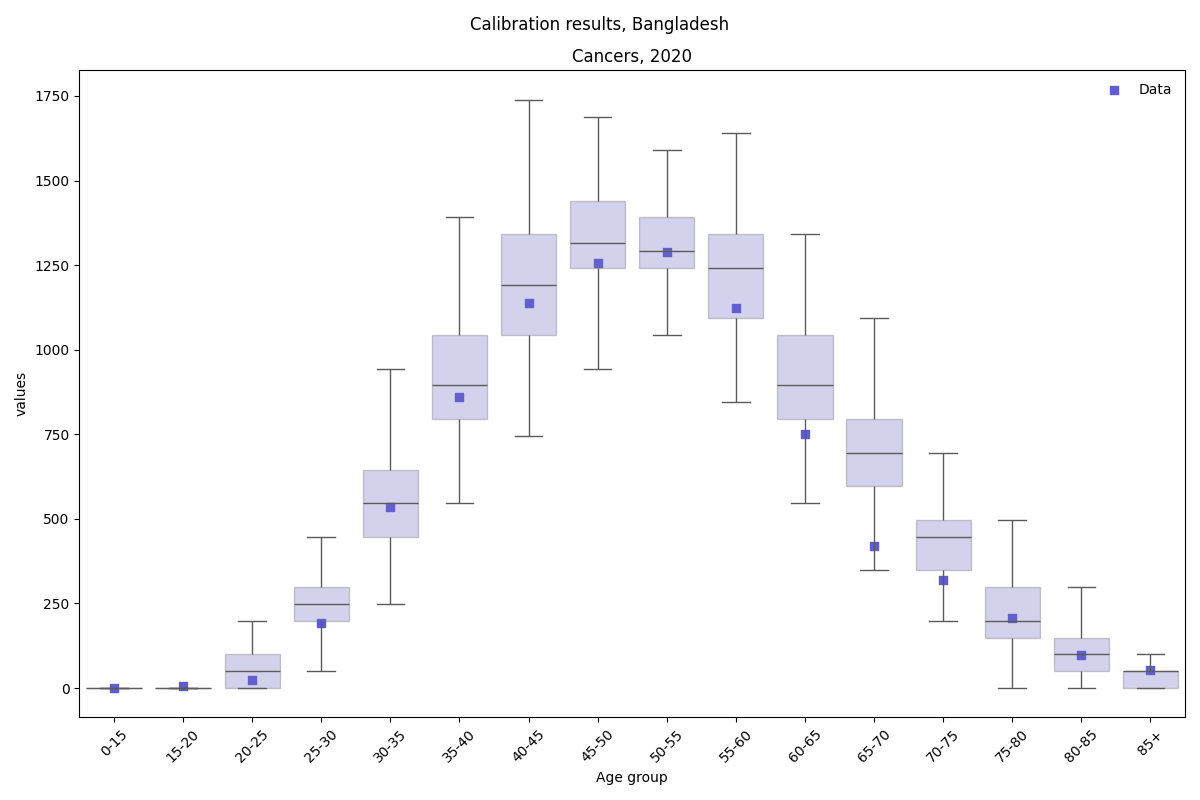

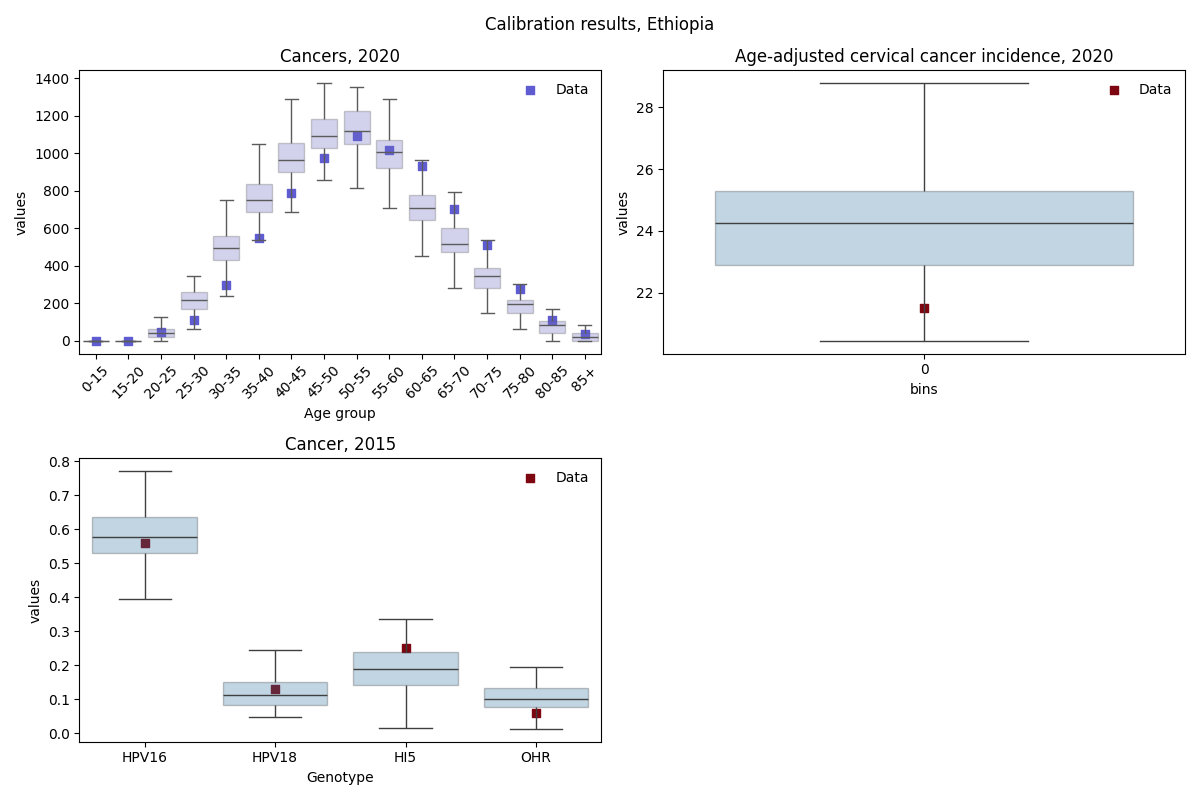

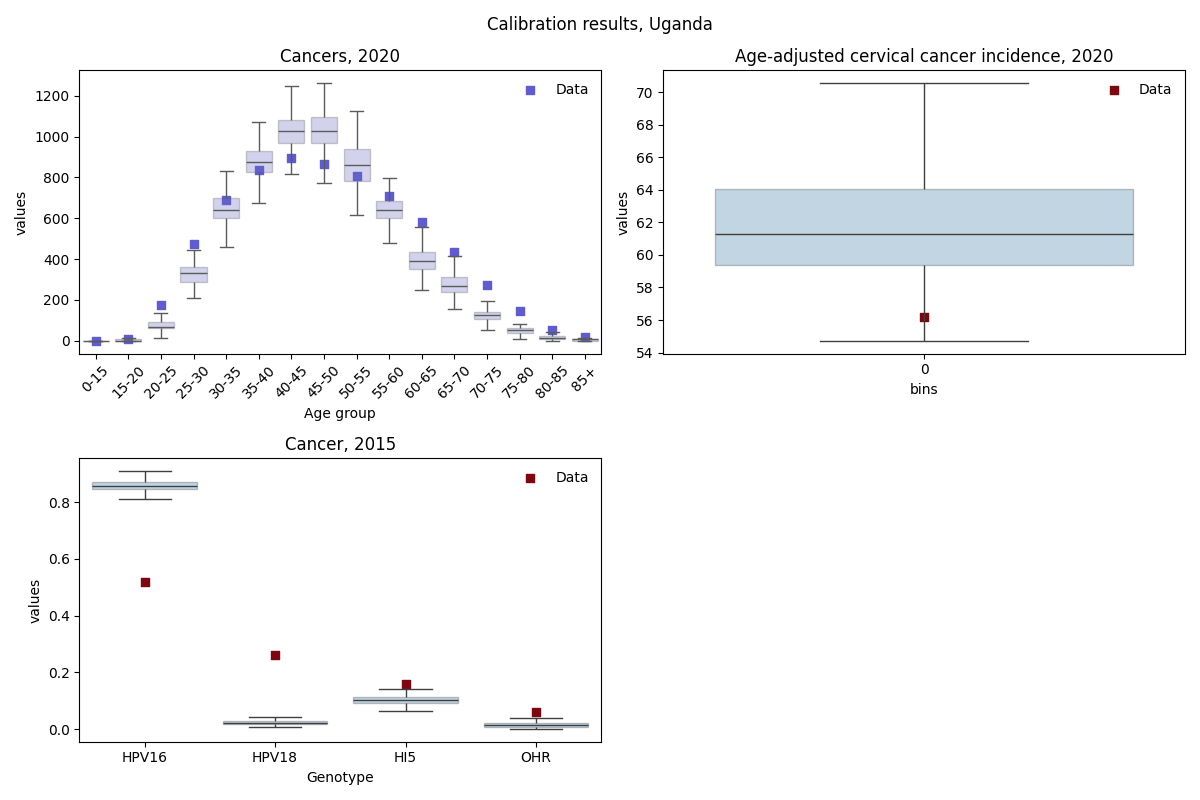

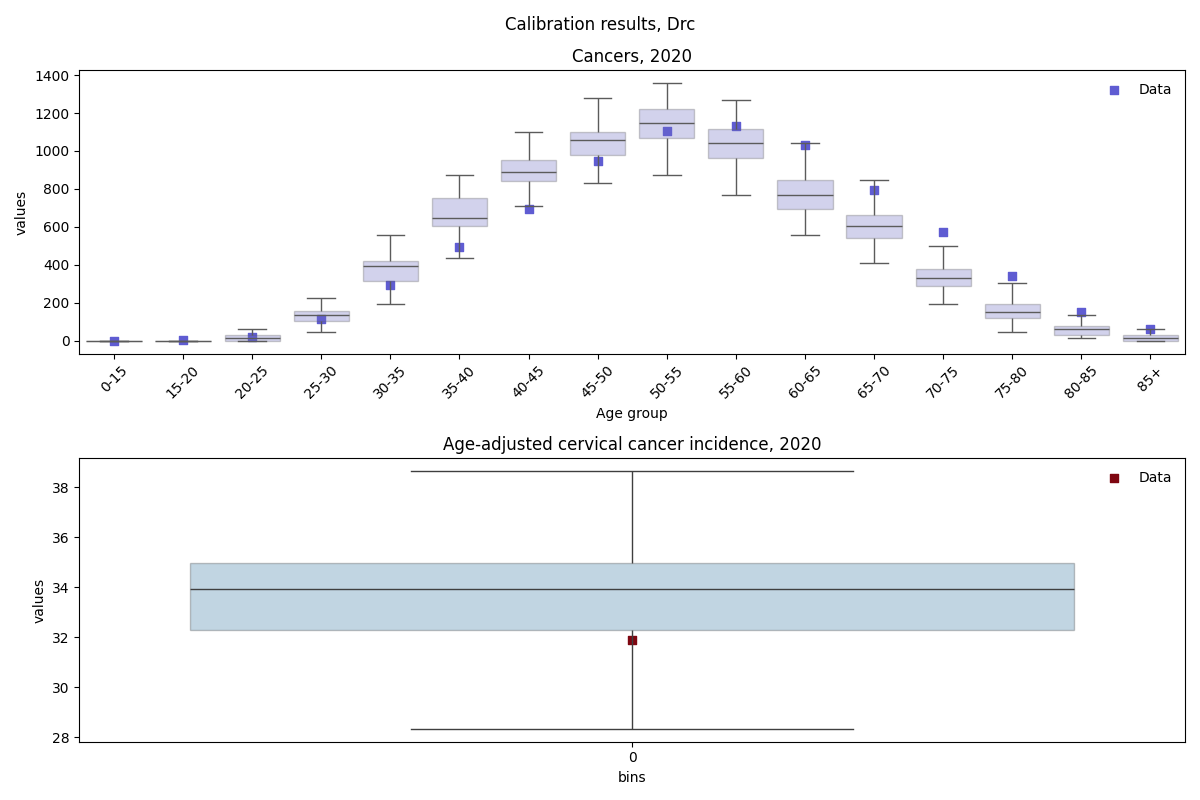

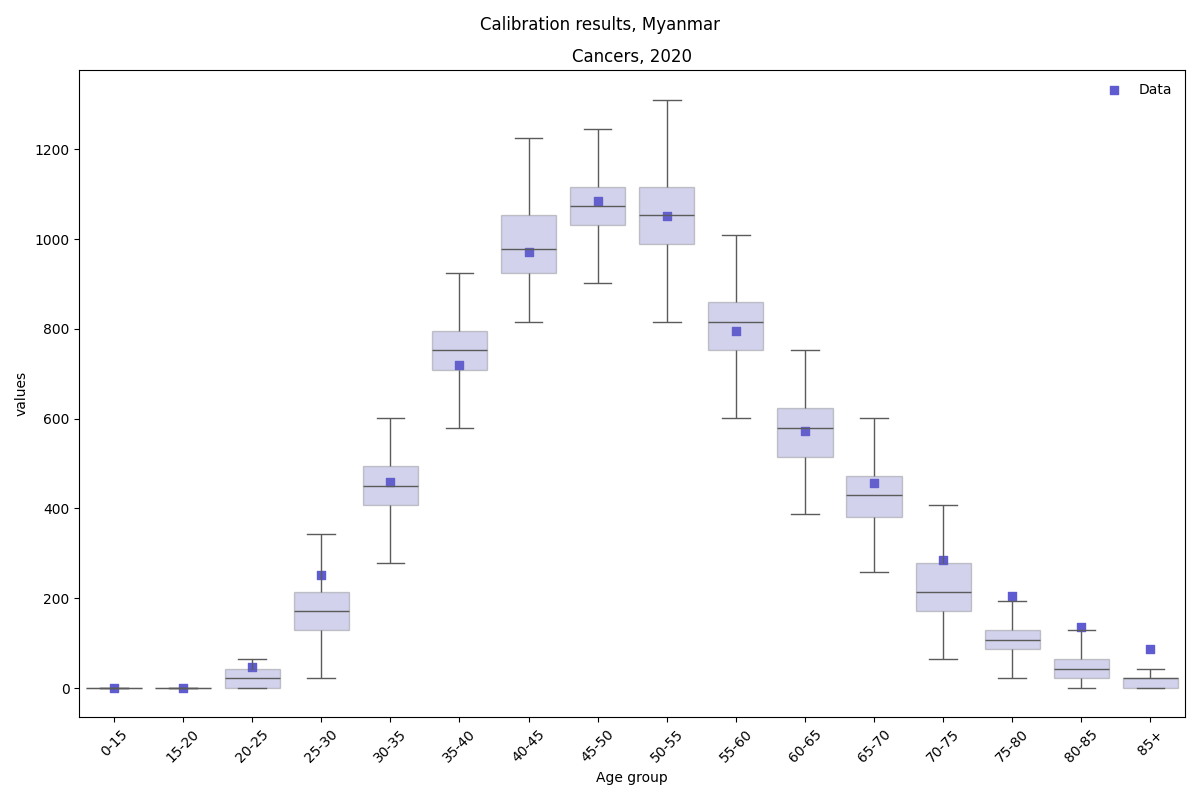

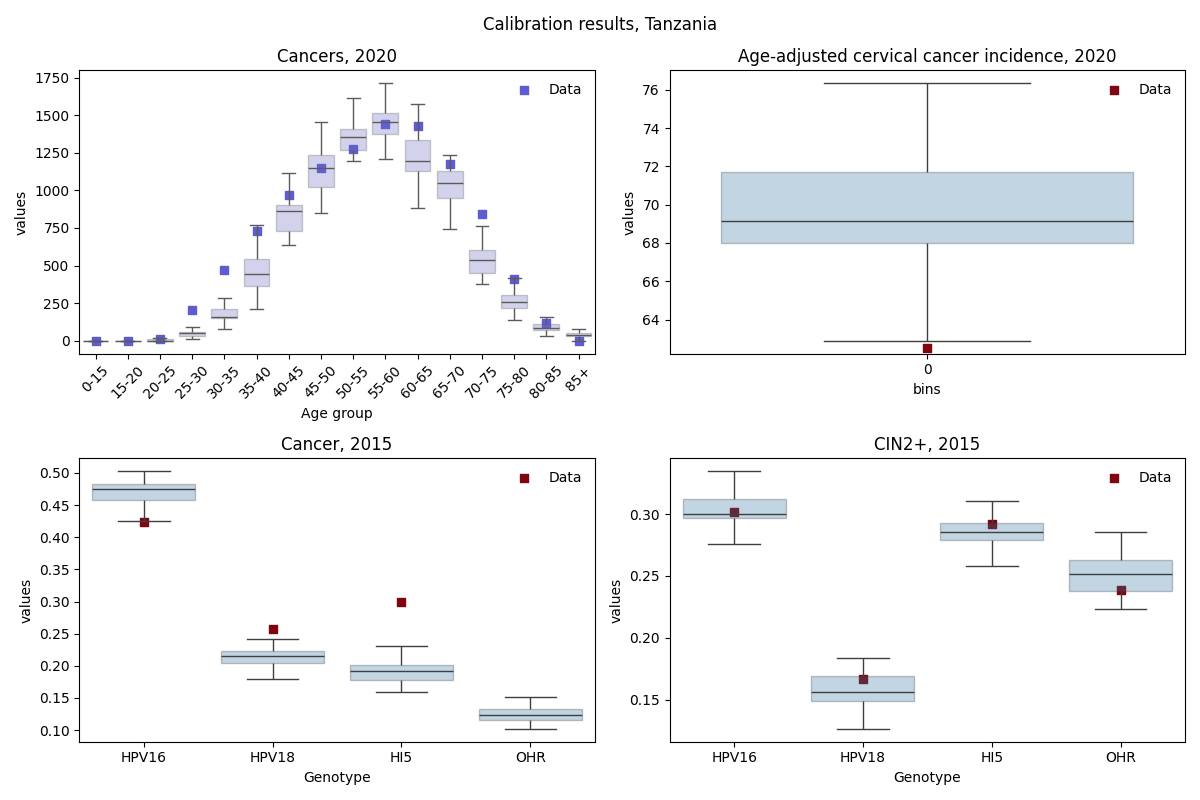

**Figure A2-10.** Calibration results for each country. Model output for the top 100 parameter sets is plotted alongside corresponding target data.

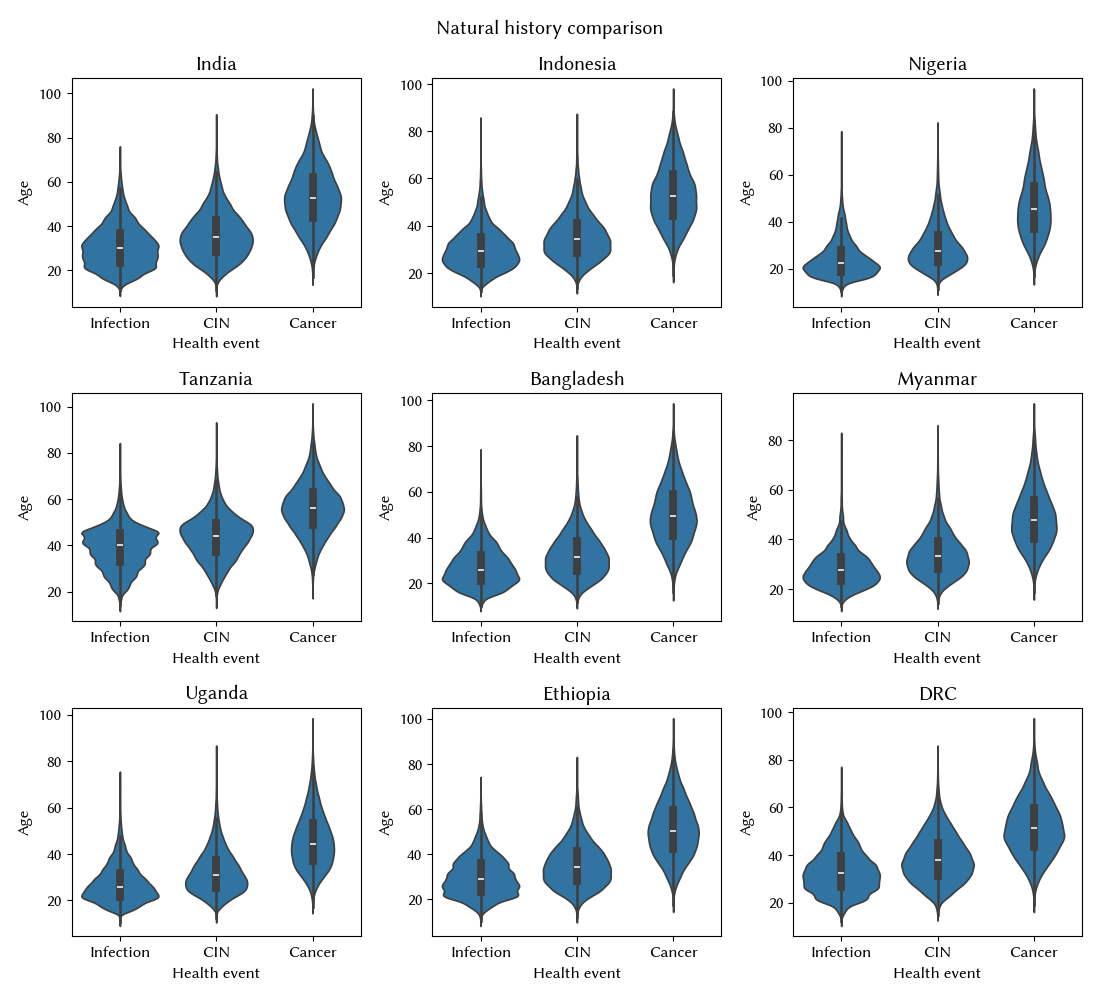

**Figure A11.** Estimated natural history of causal HPV infections implied by the calibrations for all 9 countries.

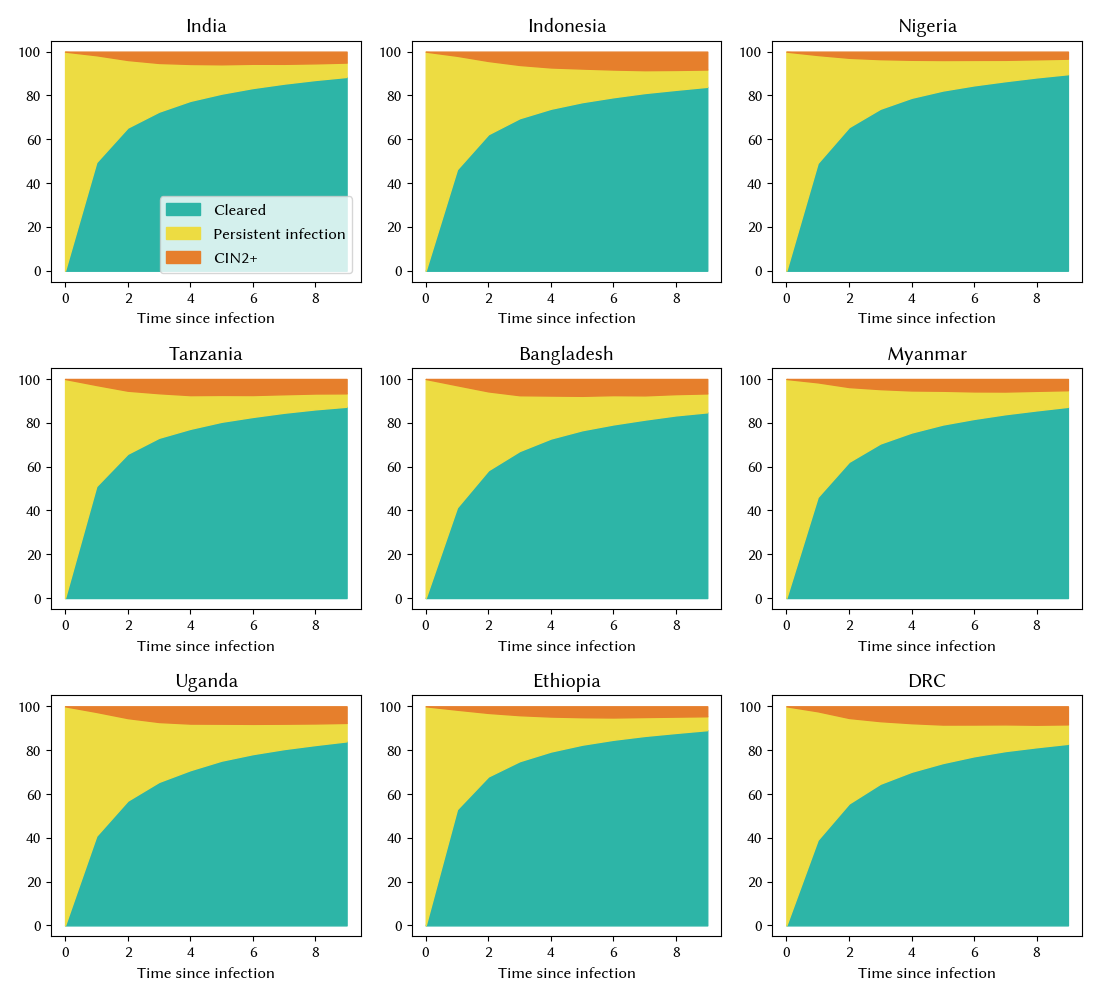

**Figure A12.** Estimated distribution of infection outcomes for all HPV infections over time implied by the calibrations for all 9 countries.

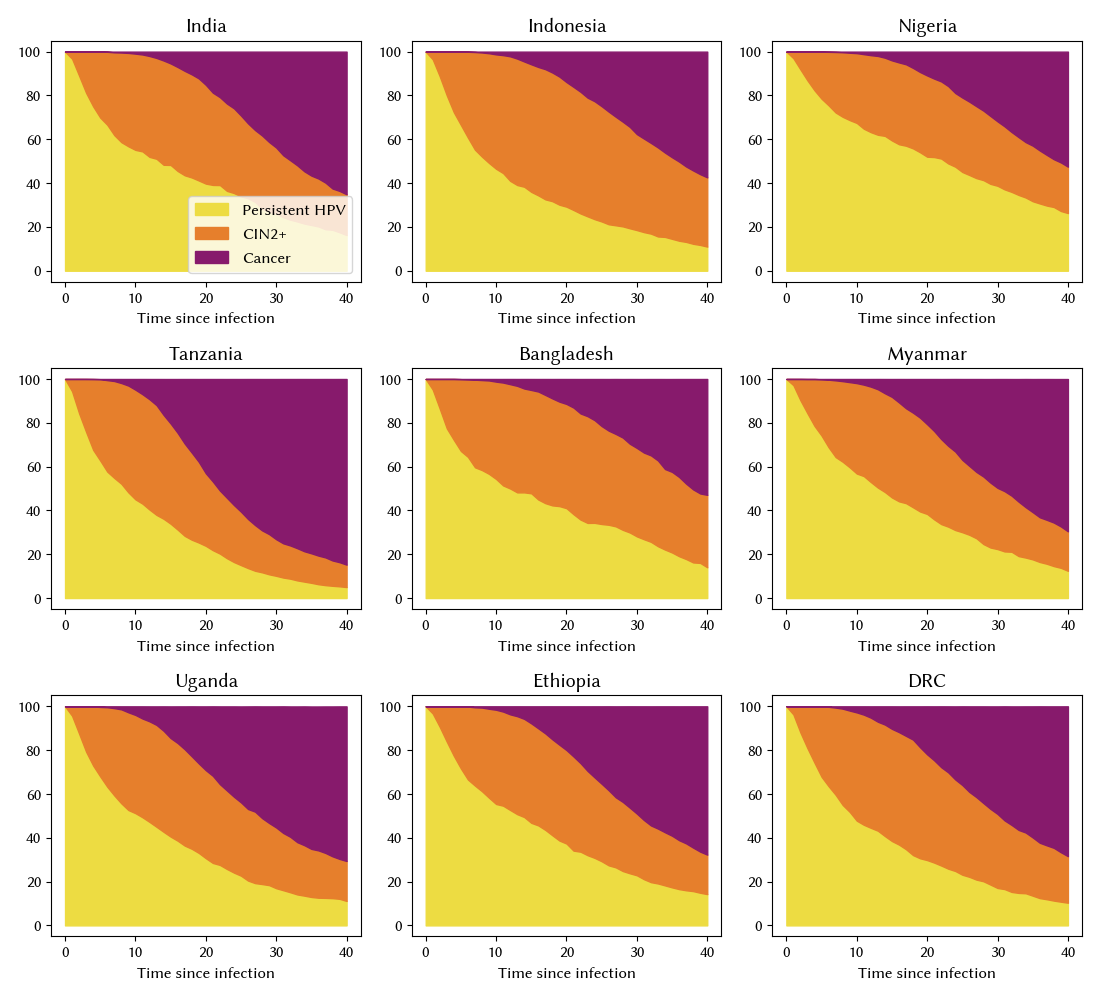

**Figure A13.** Estimated distribution of outcomes for persistent infections over time implied by the calibrations for all 9 countries.

#### 1.3 Calculation of disability adjusted life years (DALYs)

DALYs were calculated as the sum of years of life lived with disability (YLD) and years of life lost (YLD) based upon the following equation:

$DALYs= \sum_{i}^{Y} years with cancer*\mathrm{disutility}_{\mathrm{cancer}}+LE_{standard}-age of cancer death$, (3)

where $\mathrm{disutility}_{\mathrm{cancer}}$ was set to 0.29 and $LE_{standard}$ set to 88.8 years.

#### 1.4 Data and materials availability

The source code for HPVsim, including documentation and tutorials, is freely available via [GitHub](http://hpvsim.org). This manuscript refers to all features and parameters of version 2.0.0 of HPVsim. Any subsequent changes will be documented on [GitHub](http://hpvsim.org). The analysis repository can also be found on GitHub.

### 2 Additional Model Results

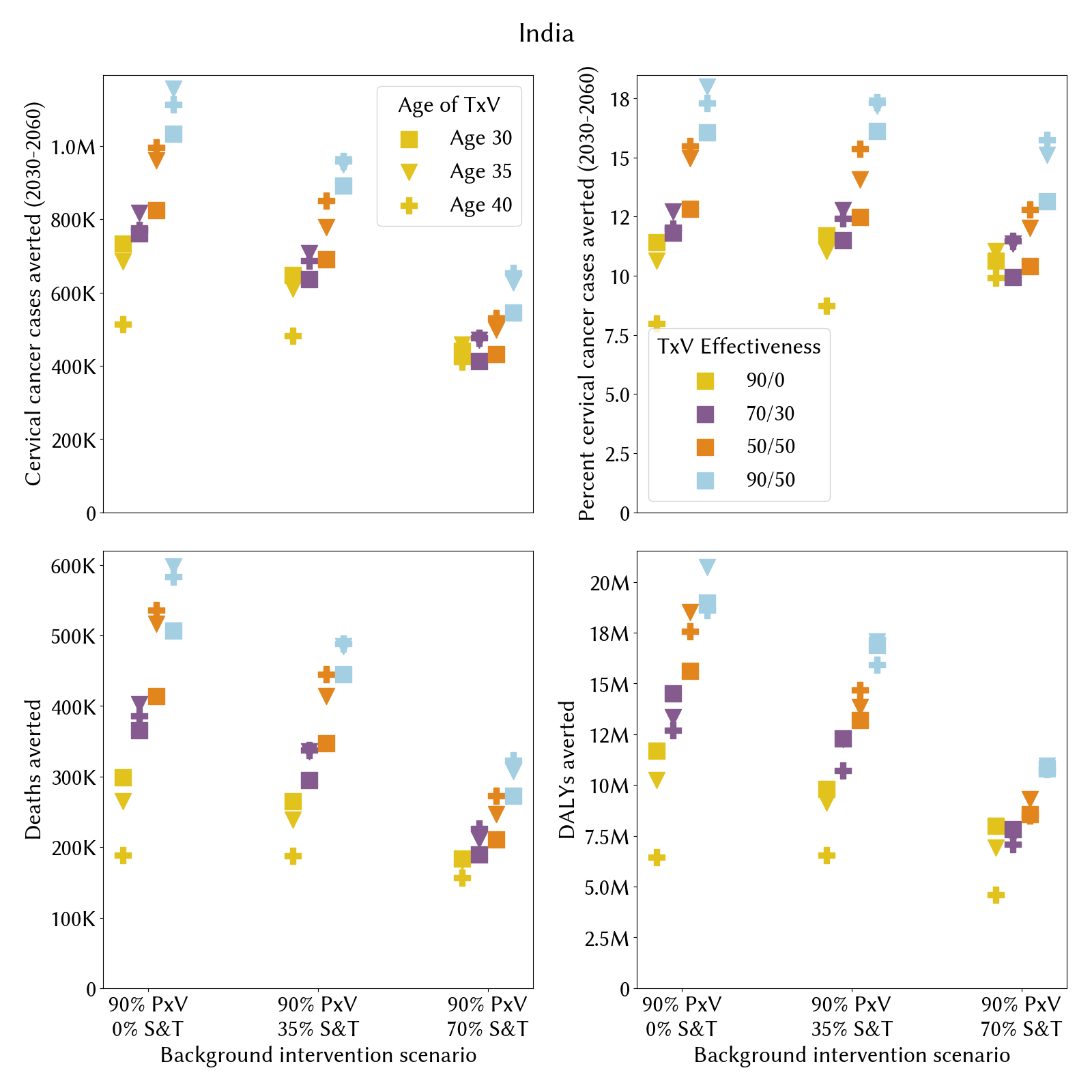

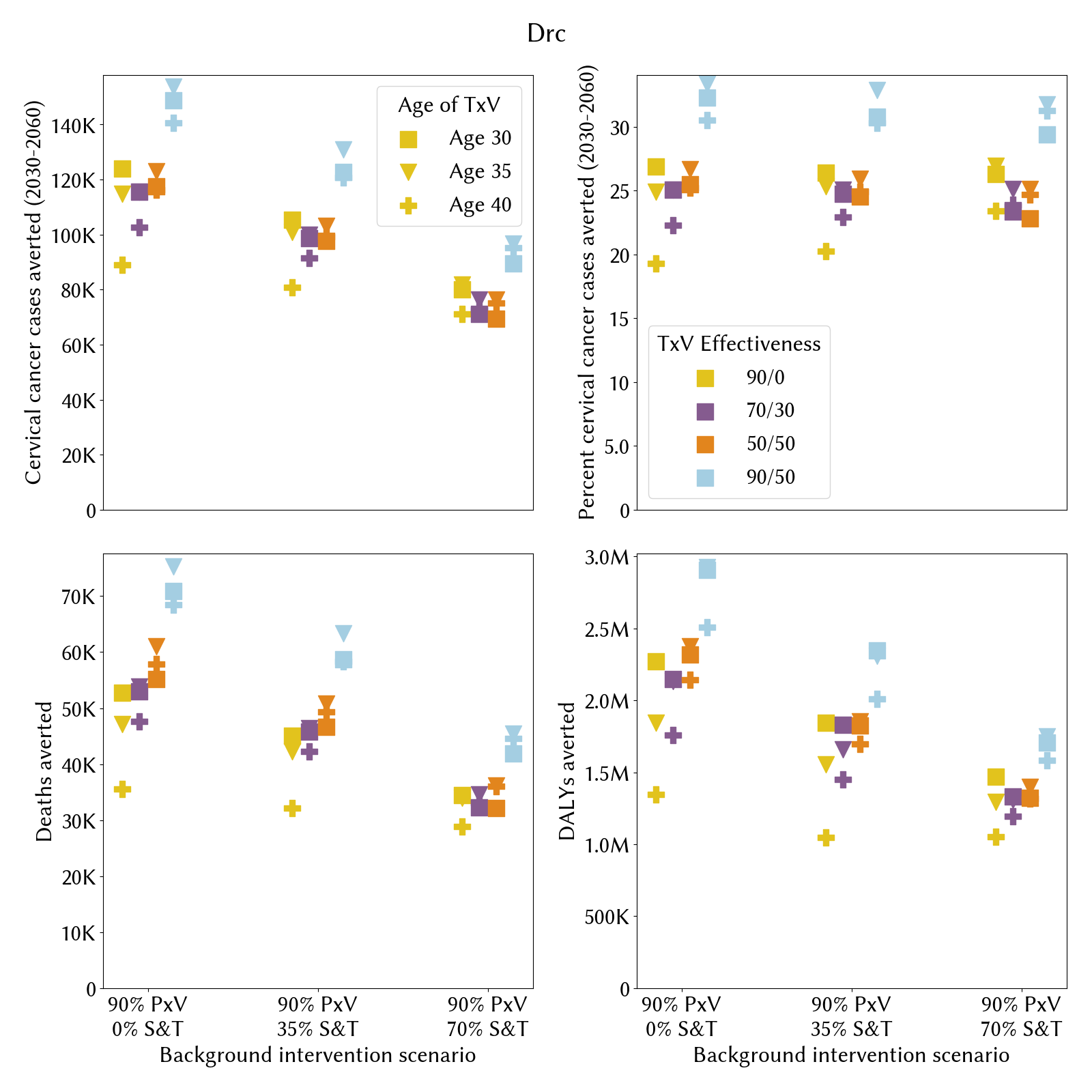

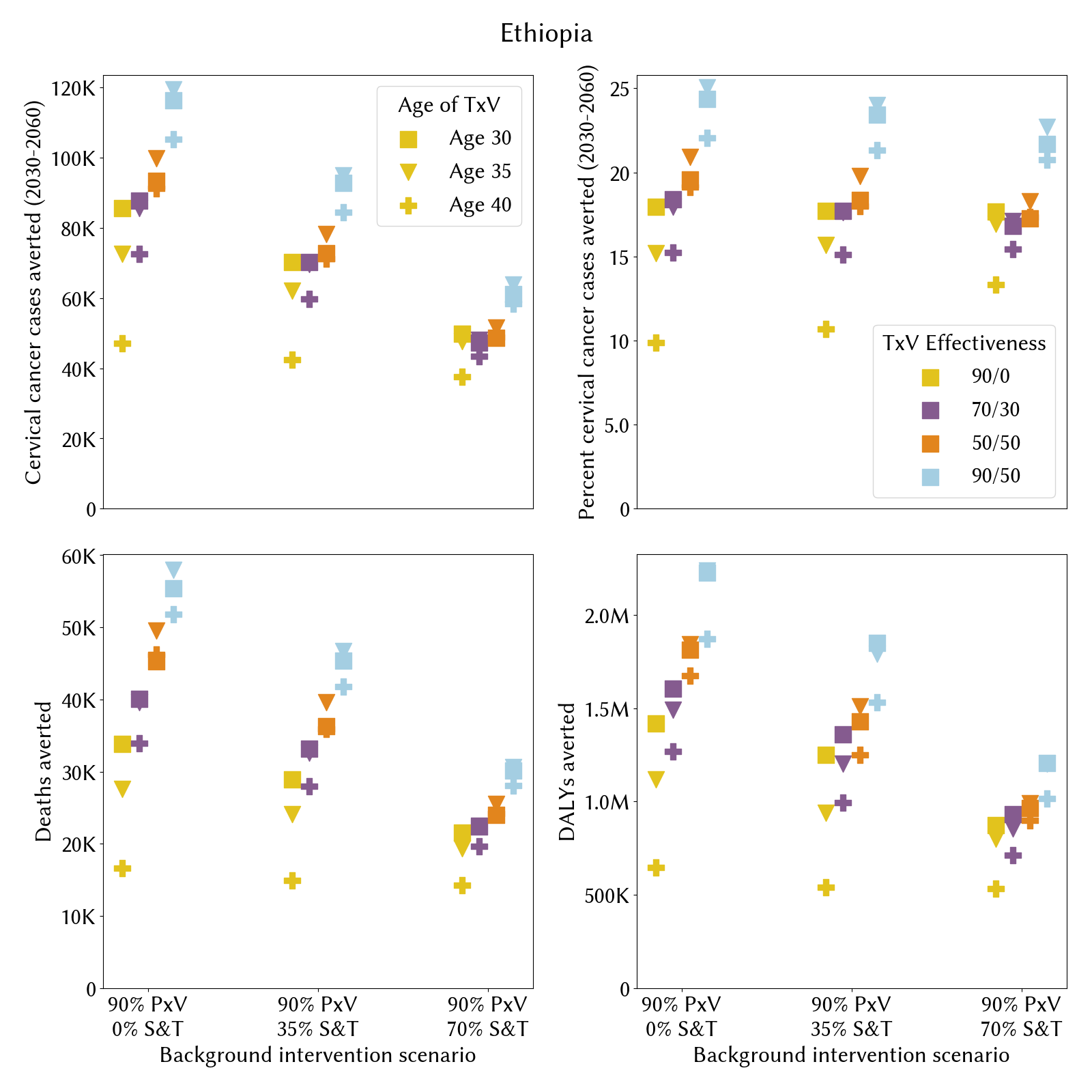

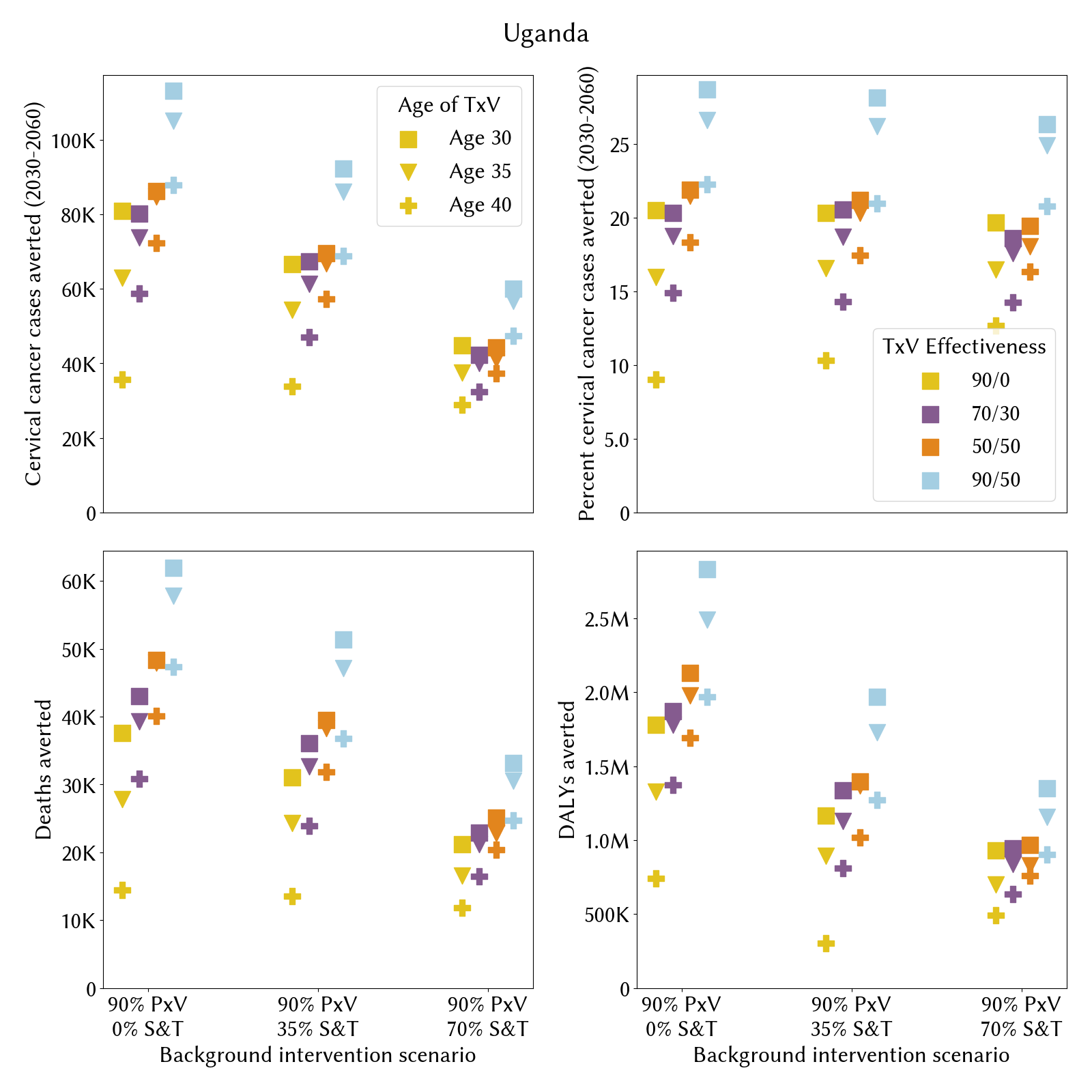

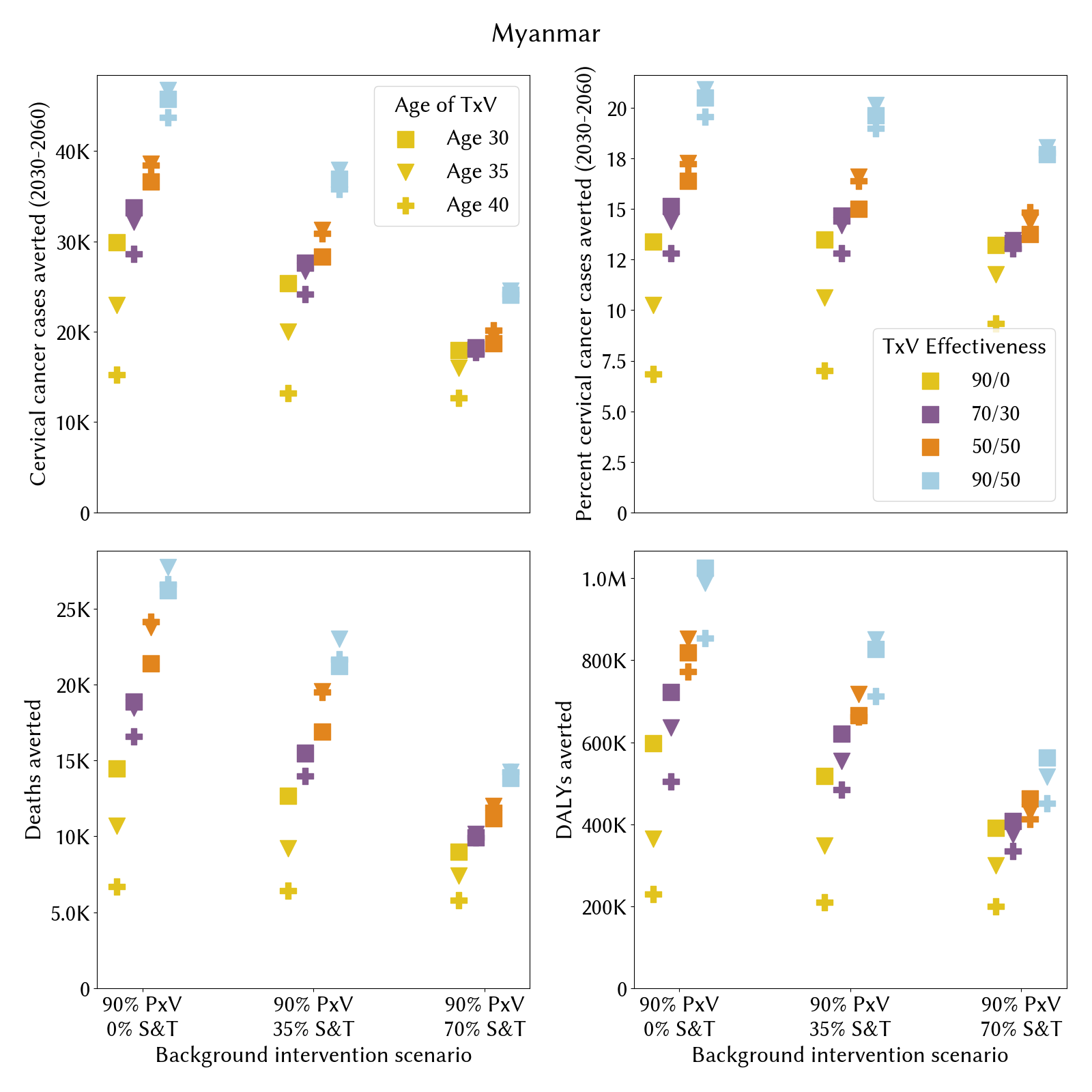

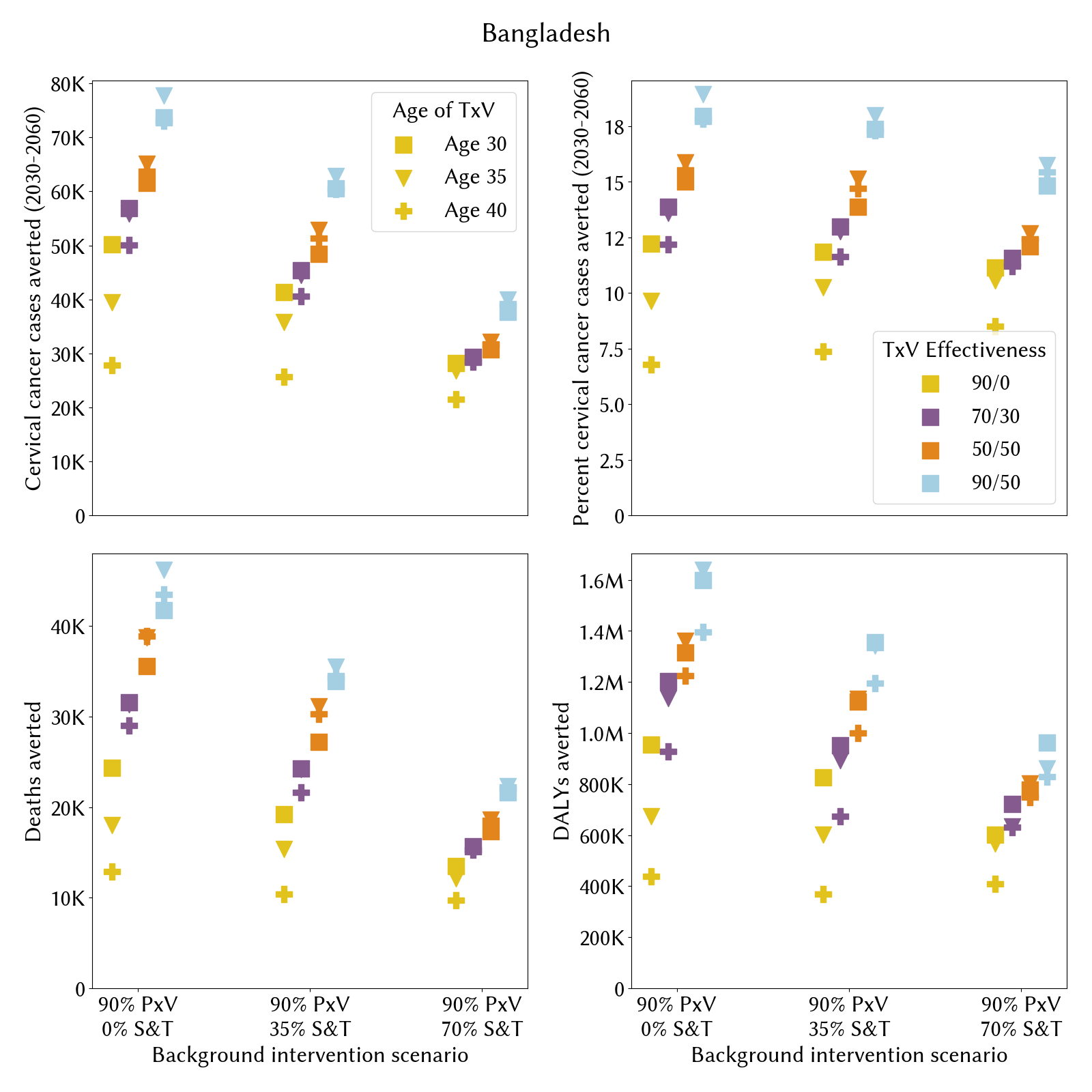

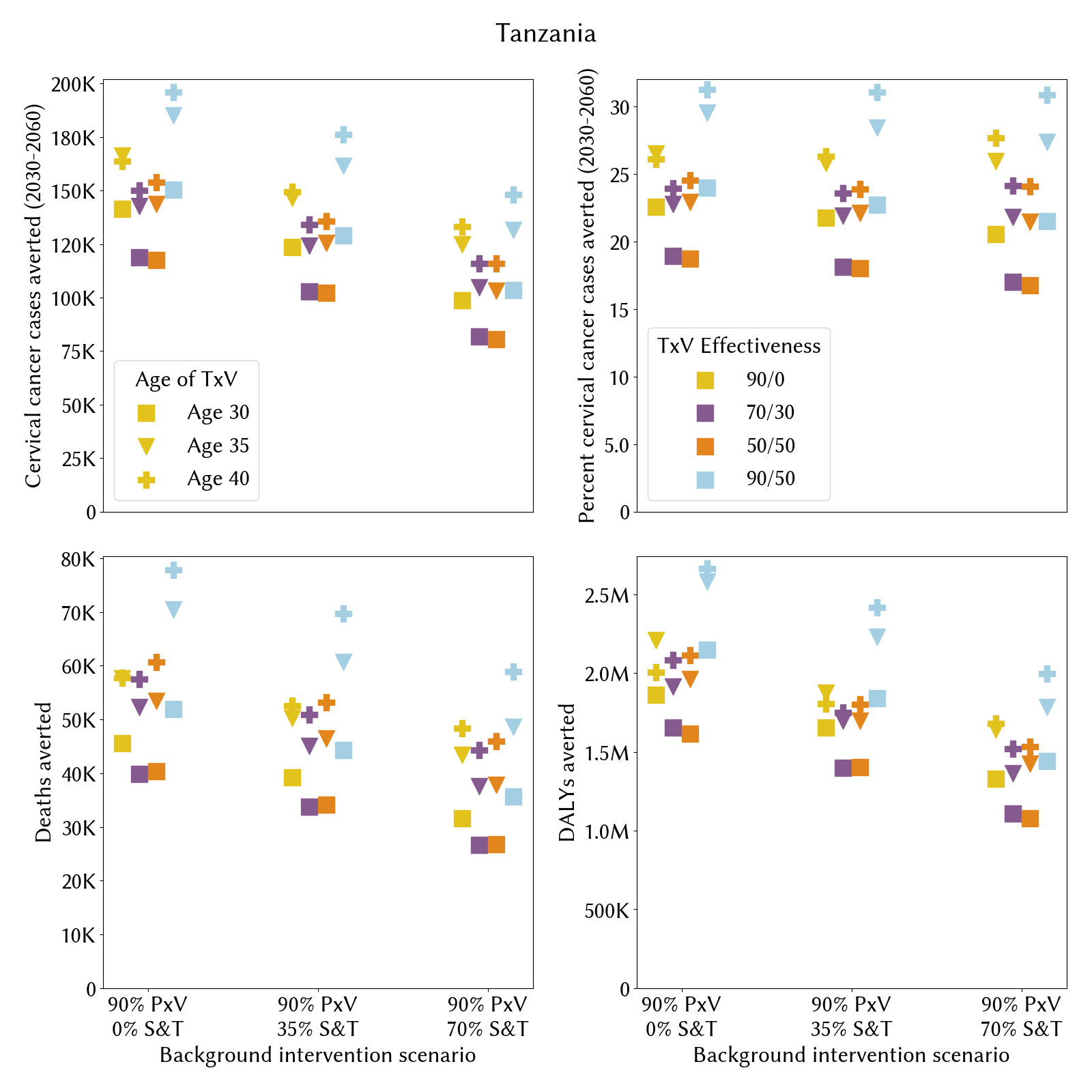

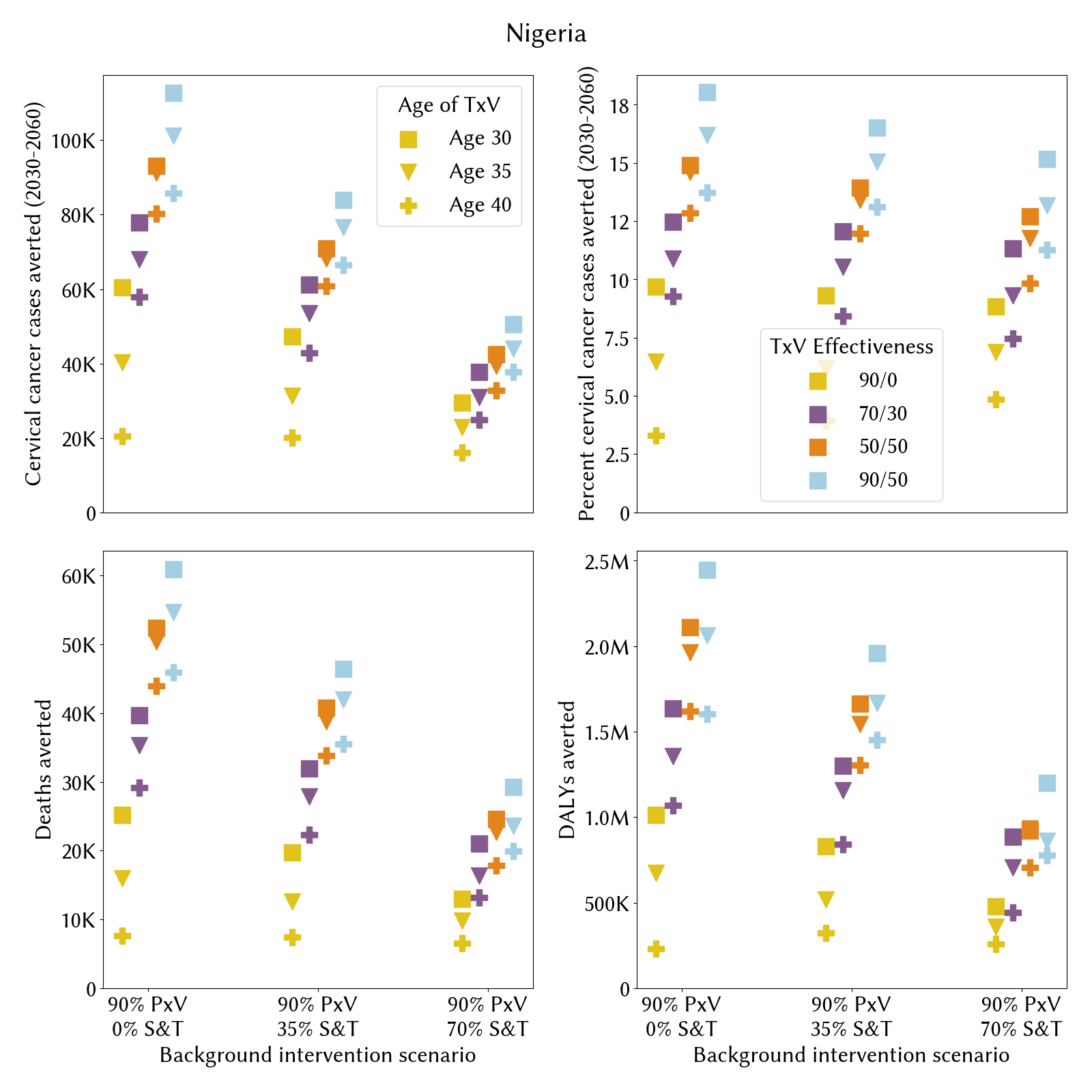

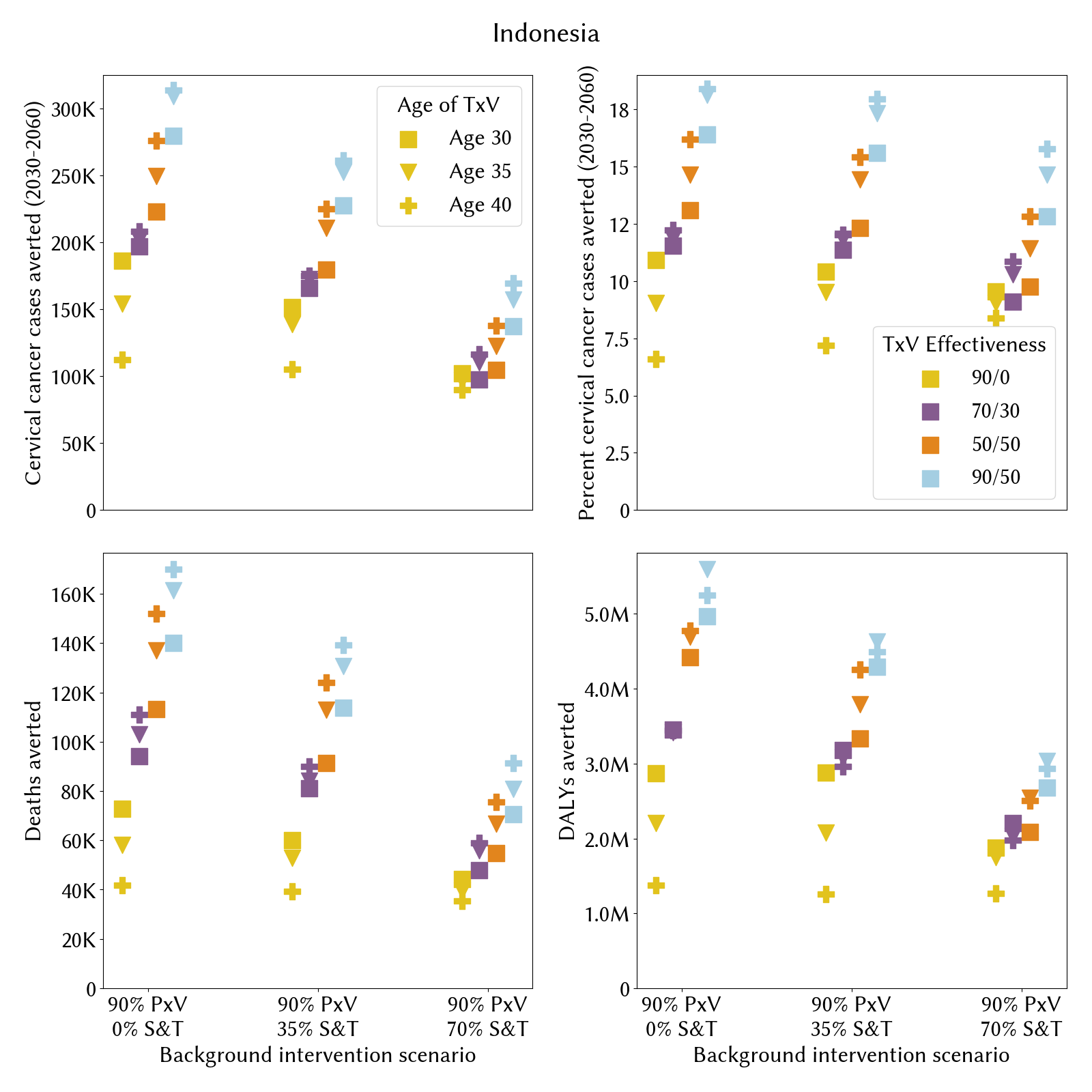
**Figure A14-23.** Therapeutic vaccine impact for each country, varying the TxV effectiveness, background intervention scale-up, and age of administration.
